# Coverage of Empirical Confidence Intervals for the Ensemble Mean of General Circulation Models in Projecting Climate Change Impacts on Health

**DOI:** 10.64898/2026.08.01.26359345

**Authors:** Yui Tomo

## Abstract

In projections of climate change impacts on health, the mean impact across general circulation models (GCMs) is commonly reported with an empirical confidence interval (eCI) formed from quantiles of impacts pooled across GCMs and coefficient draws. We specify the estimands and predictands and examine intervals aligned with them. The eCI is aligned with the impact under a GCM selected uniformly at random from the considered set, and its nominal level does not indicate its coverage for the mean. We examine coverage as the observed-data sample size increases, with a fixed number of GCMs and infinitely many coefficient draws. At the 95% nominal level, coverage for the mean over the considered GCMs converges to one with fewer than 40 GCMs unless their impacts are all equal, and can converge to zero with more than 40. When GCMs are sampled independently from a population with a normal distribution of impacts, coverage for the population mean converges to 0.75 and 0.9375 for three and five GCMs, respectively. For each of the two estimands and two predictands, we identify an interval whose coverage converges to the nominal level under the respective assumptions. In simulated heat-related mortality projections, coverage of the 95% eCI for the mean over the considered GCMs was at least 0.99 in 22 of 26 settings. A London reanalysis illustrates the differences between the intervals.

## 1 Introduction

Future health impacts of climate change are commonly projected by combining an exposure– response function estimated from observed data with future exposure trajectories generated by multiple general circulation models (GCMs). For each GCM, the impact of interest, such as the number of heat-related deaths in a future period, is projected by applying the estimated exposure–response function to the exposure trajectory of the GCM, and the GCM-specific impacts are aggregated into their arithmetic mean, which we refer to as the GCM-ensemble mean. The estimation uncertainty of the exposure–response function is propagated by drawing coefficients from an approximate sampling distribution of the estimator and computing the impact under each GCM with each draw. The empirical confidence interval (eCI) is then defined as the interval between the empirical quantiles of the impacts pooled across the GCMs and the coefficient draws (Vicedo-Cabrera et al., 2019; Quijal-Zamorano et al., 2026), and it is reported together with the GCM-ensemble mean. This procedure has been used in multicountry projections of temperature- and heatwave-related mortality and in projections of temperature-related cardiovascular hospitalizations (Gasparrini et al., 2017; Guo et al., 2018; Yang et al., 2021; Zhan et al., 2023; Chen et al., 2024).

In the eCI, both the variation among the GCMs and the estimation uncertainty of the exposure–response function are summarized. However, the eCI is constructed from the quantiles of the impacts pooled across the GCMs, whereas the reported point estimate is their mean. Therefore, the nominal level of the eCI does not imply the same coverage probability for the GCM-ensemble mean, and it is not clear which quantity the eCI covers with the nominal probability. The variation among the GCMs also raises the question of whether the target of the inference is the mean over the GCMs considered in the study or the mean over a population of GCMs from which the GCMs considered in the study are regarded as a sample. This is because the multi-model ensembles in climate science are not designed as a random sample of models (Tebaldi and Knutti, 2007; Knutti et al., 2013; Abramowitz et al., 2019).

In this study, we specify the targets for which the GCM-ensemble mean is reported and the intervals aligned with each of them, and we evaluate the coverage of the eCI for these targets. We consider four targets: two estimands (the mean impact over the considered GCMs and the mean impact over a population of GCMs) and two predictands (the impact under a GCM selected uniformly at random from the considered set and the impact under a new GCM). In terms of these targets, we clarify that the eCI used in previous studies is an interval aligned with the first predictand, and we show that its coverage for the two estimands can deviate from the nominal level. We then examine the intervals in a simulation study mimicking projections of heat-related mortality and in a reanalysis of the projections in London in the tutorial of Quijal-Zamorano et al. (2026).

## 2 Targets and intervals

### 2.1 Problem setup and procedures in previous studies

Let *θ* ∈ ℝ^*p*^ denote the coefficient vector of the exposure–response function, let *θ*_0_ denote its true value, and let 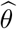 and 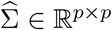 denote its estimator and the estimated covariance matrix of 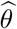. For *l* = 1, …, *L*, let *H*_*l*_ : ℝ^*p*^ → ℝ denote the map from the coefficient vector to the impact projected under GCM *l*, where *L* is the number of GCMs. For example, *H*_*l*_(*θ*) is the number of heat-related deaths in a future period computed from the temperature trajectory of GCM *l* and the exposure–response function with the coefficient vector *θ*. The GCM-ensemble mean and its point estimate are

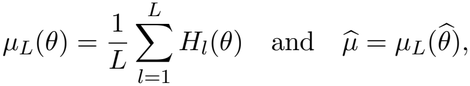

respectively.

To propagate the estimation uncertainty, coefficient vectors *θ*^(1)^, …, *θ*^(*J*)^ are drawn independently from the multivariate normal distribution 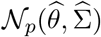. Let *µ*^(*j*)^ = *µ*_*L*_(*θ*^(*j*)^) (*j* = 1, …, *J*). For 0 *< u <* 1 and a finite multiset *S* of real numbers, let *Q*_*u*_(*S*) denote the empirical *u*-quantile of *S*. For a nominal level 1 − *α* with 0 *< α <* 1, the eCI is given by

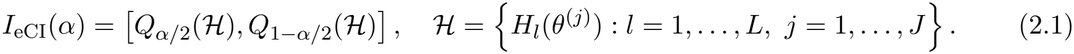

This point estimate and the eCI are used in previous studies and tutorials in the field (Vicedo-Cabrera et al., 2019; Quijal-Zamorano et al., 2026), in which 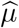 is reported together with *I*_eCI_(*α*).

### 2.2 Estimands and predictands

To examine the alignment between the point estimate and the interval, we consider four targets.

We first introduce two estimands. The first is the mean over the considered GCMs, *µ*_*L*_(*θ*_0_), for which the maps *H*_1_, …, *H*_*L*_ are fixed and the randomness arises only from the observed data used for the estimation of *θ*.

The second is the mean over a population of GCMs. Suppose that the maps *H*_1_, …, *H*_*L*_ are those of GCMs *G*_1_, …, *G*_*L*_ sampled independently from a population of GCMs and independently of the observed data, and let *H*_*G*_ denote the map of a GCM *G* sampled from the population. Then, the estimand is *µ*_∞_(*θ*_0_) = E*{H*_*G*_(*θ*_0_)*}*.

We then introduce two predictands, i.e., random quantities to be predicted. The first is *H*_*s*_(*θ*_0_), the impact under a GCM selected uniformly at random from the considered set, where *s* is uniformly distributed on *{*1, …, *L}* and independent of the observed data.

The second is 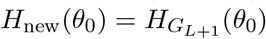, the impact under a new GCM *G*_*L*+1_ sampled from the population independently of *G*_1_, …, *G*_*L*_ and of the observed data.

### 2.3 Definitions of the confidence and prediction intervals

An interval *I* is a confidence interval for *µ*_*L*_(*θ*_0_) at the level 1 − *α* if

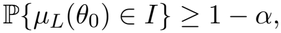

for every value of *θ*_0_ (Neyman, 1937; Lehmann and Romano, 2005), where ℙ denotes the probability under repeated sampling of the observed data. Similarly, *I* is an asymptotic confidence interval at the level 1 − *α* if the limit inferior of this probability is at least 1 − *α* for every value of *θ*_0_ as the sample size of the observed data increases. For *µ*_∞_(*θ*_0_), the probability in the definition of the confidence interval is taken over both the observed data and the sampling of the GCMs.

Also, an interval *I* is a prediction interval for *H*_*s*_(*θ*_0_) at the level 1 − *α* if

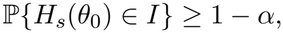

for every value of *θ*_0_, where the probability is taken over both the observed data and *s*, and an asymptotic prediction interval is defined in the same manner. For *H*_new_(*θ*_0_), the probability is taken over both the observed data and the sampling of *G*_1_, …, *G*_*L*+1_.

### 2.4 Intervals aligned with the targets

We first consider *µ*_*L*_(*θ*_0_) and examine the interval aligned with it. We introduce the interval obtained by averaging the impacts over the GCMs before taking the quantiles,

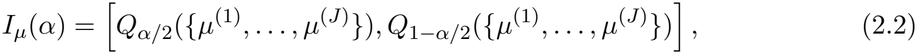

in which the estimation uncertainty is propagated through the same functional *µ*_*L*_ as the point estimate 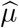. Let 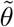 follow 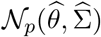 given the data. Then, under a regularity condition, as *J* → ∞, the endpoints of *I*_*µ*_(*α*) converge to the *α/*2- and (1 − *α/*2)-quantiles of the conditional distribution of 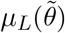. See Section S2 of the Supplementary Material for the condition and the proof. Therefore, *I*_*µ*_(*α*) is the simulation-based interval for a function of the estimated coefficients (Krinsky and Robb, 1986), which coincides with the delta-method confidence interval when *µ*_*L*_ is linear.

On the other hand, we examine the target with which *I*_eCI_(*α*) is aligned. Let *s* be uniformly distributed on *{*1, …, *L}* and independent of 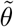. Then, under a regularity condition, as *J* → ∞, the endpoints of *I*_eCI_(*α*) converge to the *α/*2- and (1 − *α/*2)-quantiles of the conditional distribution of 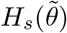, i.e., of the equally weighted mixture of the conditional distributions of 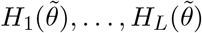. See Section S2 of the Supplementary Material for the condition and the proof. In this sense, *I*_eCI_(*α*) is an interval aligned with the impact under a GCM selected uniformly at random from the considered set.

For *µ*_∞_(*θ*_0_), we decompose the error of the point estimate as

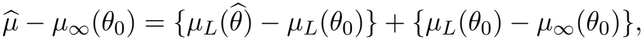

where the first term arises from the estimation of *θ* and the second from the sampling of the GCMs. Given the GCMs, the first term has mean approximately zero and the second term is fixed, so that the two terms are approximately uncorrelated, and we have 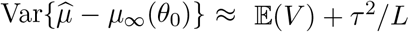, where *V* denotes the variance of 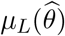 given the GCMs, *τ* ^2^ denotes the variance of *H*_*G*_(*θ*_0_) over the population, and the expectation is taken over the sampling of the GCMs. Here, *τ* ^2^*/L* is the variance of *µ*_*L*_(*θ*_0_) for the GCMs sampled independently. We estimate *V* and *τ* ^2^ by

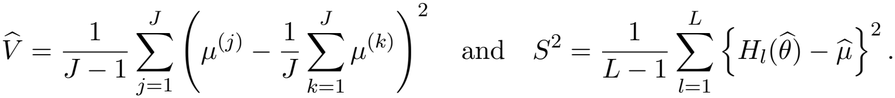

Note that *S*^2^ also includes the differences among 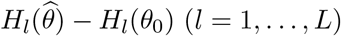, which vanish in the location-shift model in Section 3.3. For *ν >* 0 and 0 *< u <* 1, let *t*_*ν,u*_ denote the *u*-quantile of the *t* distribution with *ν* degrees of freedom. We define the interval for *µ*_∞_(*θ*_0_) as

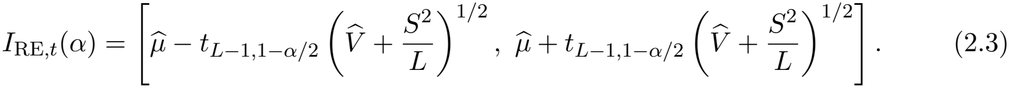

In *I*_RE,*t*_(*α*), we use the quantile of the *t* distribution with *L* − 1 degrees of freedom to account for the estimation of *τ* ^2^ from *L* GCMs, as in the confidence interval derived from the test of Hartung and Knapp (2001) for the overall effect in random-effects meta-analysis.

As a simulation-based variant within the same procedure as the eCI, we also consider the interval *I*_RE,sim_(*α*) between the empirical *α/*2- and (1 − *α/*2)-quantiles of *µ*^(*j*)^ + (*S/L*^1*/*2^)*U* ^(*j*)^ (*j* = 1, …, *J*), where *U* ^(1)^, …, *U* ^(*J*)^ are drawn independently from the *t* distribution with *L* − 1 degrees of freedom. For the predictand *H*_new_(*θ*_0_), as in the prediction interval in random-effects meta-analysis (Higgins et al., 2009), we add the variance of the impact under a new GCM and define the prediction interval

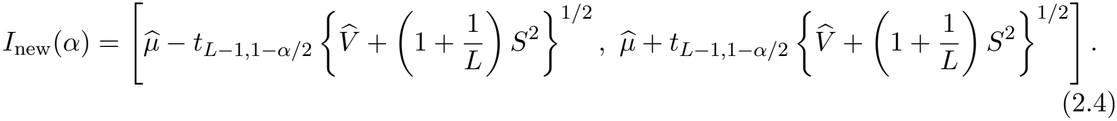

Hereafter, for the theoretical results, we use the notation *I*_eCI_(*α*), *I*_*µ*_(*α*), *I*_RE,*t*_(*α*), *I*_RE,sim_(*α*), and *I*_new_(*α*) for the intervals in which the empirical quantiles and the sample variance over the draws are replaced by the quantiles and the variance of the conditional distributions given the data, where the *u*-quantile of a distribution function *F* is inf*{x* : *F* (*x*) ≥ *u}*. Under the conditions in Section S2 of the Supplementary Material and if the conditional second moment of 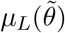 given the data is finite, these intervals are the limits of the intervals with *J* coefficient draws as *J* → ∞.

Table 1 summarizes the targets and the intervals aligned with them, where the point estimate is 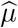 for all targets.

**Table 1:**
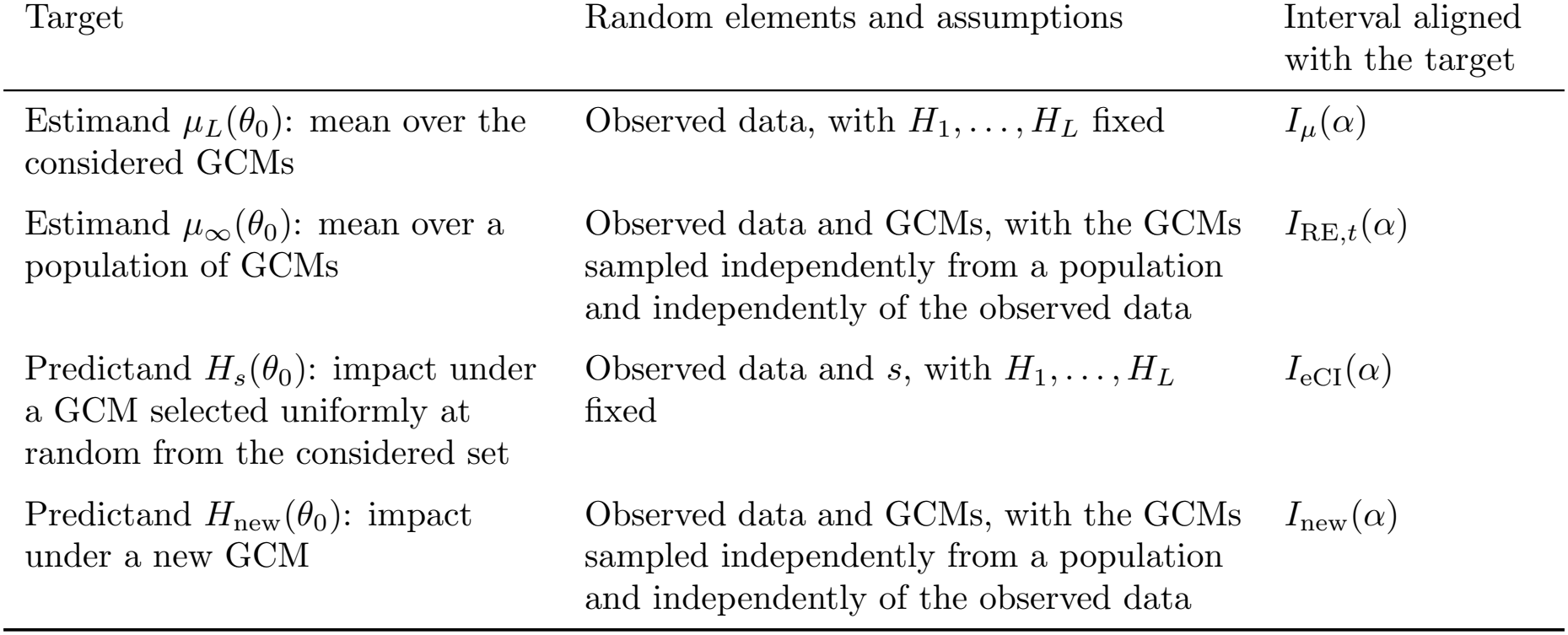
Targets for which the GCM-ensemble mean 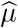 is reported and the intervals aligned with them.

| Target | Random elements and assumptions | Interval aligned with the target |
|---|---|---|
| Estimand μ<sub>L</sub>(θ<sub>0</sub>): mean over the considered GCMs | Observed data, with H<sub>1</sub>, …, H<sub>L</sub> fixed | I<sub>μ</sub>(α) |
| Estimand μ<sub>∞</sub>(θ<sub>0</sub>): mean over a population of GCMs | Observed data and GCMs, with the GCMs sampled independently from a population and independently of the observed data | I<sub>RE,t</sub>(α) |
| Predictand H<sub>s</sub>(θ<sub>0</sub>): impact under a GCM selected uniformly at random from the considered set | Observed data and s, with H<sub>1</sub>, …, H<sub>L</sub> fixed | I<sub>eCI</sub>(α) |
| Predictand H<sub>new</sub>(θ<sub>0</sub>): impact under a new GCM | Observed data and GCMs, with the GCMs sampled independently from a population and independently of the observed data | I<sub>new</sub>(α) |

## 3 Coverage of the intervals for the targets

In this section, we evaluate the coverage of the intervals in Table 1 for their targets and that of *I*_eCI_(*α*) for the two estimands.

### 3.1 Mean over the considered general circulation models

We first consider the limit as the sample size *n* of the observed data used for the estimation of *θ* increases with the GCMs fixed. As defined in Section 2.4, the intervals are those with *J* → ∞, and 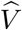 is the conditional variance of 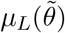 given the data. Let *H*_(1)_(*θ*_0_) ≤ *· · ·* ≤ *H*_(*L*)_(*θ*_0_) denote the order statistics of *H*_1_(*θ*_0_), …, *H*_*L*_(*θ*_0_). We consider the following assumption, which holds under standard regularity conditions for the estimators of the regression models used for the exposure–response function.

#### Assumption 1.

*As n* → ∞, 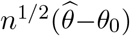 *converges in distribution to N*_*p*_(0, Σ_0_), *where* Σ_0_ ∈ ℝ^*p×p*^ *is a positive definite matrix, and* 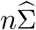 *converges in probability to* Σ_0_.

Then, for the coverage of *I*_eCI_(*α*) for *µ*_*L*_(*θ*_0_), we have the following theorem.

#### Theorem 3.1.

*Suppose that Assumption 1 is satisfied and that H*_1_, …, *H*_*L*_ *are continuous at θ*_0_. *Let k*_1_ = ⌈*Lα/*2⌉ *and k*_2_ = ⌊*Lα/*2⌋ + 1. *Then, as n* → ∞, *the following two statements hold*.

i. *If* 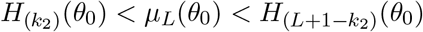, *then* ℙ*{µ*_*L*_(*θ*_0_) ∈ *I*_eCI_(*α*)*}* → 1.
ii. *If* 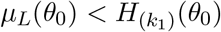 *or* 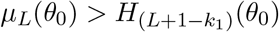, *then* ℙ*{µ*_*L*_(*θ*_0_) ∈ *I*_eCI_(*α*)*}* → 0.

See Section S3.2 of the Supplementary Material for the proof. Intuitively, as *n* increases, the impacts *H*_*l*_(*θ*^(*j*)^) over the draws concentrate around *H*_*l*_(*θ*_0_) for each GCM, so that the pooled impacts in *ℋ* concentrate on the *L* values *H*_1_(*θ*_0_), …, *H*_*L*_(*θ*_0_), each with weight 1*/L*. Then, *I*_eCI_(*α*) approaches the range of these *L* values after approximately *Lα/*2 GCMs are excluded from each end. The width of this range is determined by the differences among the GCM-specific impacts and need not shrink as *n* increases. Therefore, the coverage of *I*_eCI_(*α*) for *µ*_*L*_(*θ*_0_) converges to 1 or 0 depending on whether *µ*_*L*_(*θ*_0_) lies inside or outside this range. This is a consequence of the construction of *I*_eCI_(*α*) as an interval aligned with *H*_*s*_(*θ*_0_).

In particular, from this theorem, if *L <* 2*/α*, then *k*_1_ = *k*_2_ = 1, and the coverage of *I*_eCI_(*α*) for *µ*_*L*_(*θ*_0_) converges to 1 unless *H*_1_(*θ*_0_) = *· · ·* = *H*_*L*_(*θ*_0_), i.e., *I*_eCI_(*α*) is conservative for *µ*_*L*_(*θ*_0_) in large samples. If *L >* 2*/α*, then *k*_1_ ≥ 2, and the coverage converges to 0 when a few GCMs with extreme impacts shift *µ*_*L*_(*θ*_0_) outside the range between the *k*_1_th smallest and the *k*_1_th largest of *H*_1_(*θ*_0_), …, *H*_*L*_(*θ*_0_), e.g., when *H*_1_(*θ*_0_) = *· · ·* = *H*_*L*−1_(*θ*_0_) *< H*_*L*_(*θ*_0_). The threshold 2*/α* is 40 for 1 − *α* = 0.95, 20 for 1 − *α* = 0.90, and approximately 5.9 for 1 − *α* = 0.66.

In addition to 1 − *α* = 0.95, we also consider 1 − *α* = 0.90 and 0.66, which are the lower bounds of the probabilities for the terms “very likely” and “likely” in the calibrated uncertainty language of the Intergovernmental Panel on Climate Change (Mastrandrea et al., 2010), and intervals at 1 − *α* = 0.66 have been reported in projections of temperature-related mortality (Lay et al., 2021).

In contrast, for the intervals aligned with *µ*_*L*_(*θ*_0_) and *H*_*s*_(*θ*_0_), we consider the following assumption in addition to Assumption 1.

#### Assumption 2.

*For l* = 1, …, *L, the map H*_*l*_ *is continuously differentiable in a neighborhood of θ*_0_, *and* ∇*H*_*l*_(*θ*_0_) *≠* 0.

Then, we have the following theorem.

#### Theorem 3.2.

*Suppose that Assumptions 1 and 2 are satisfied. Then, as n* → ∞, *the following two statements hold*.

i. *If* ∇*µ*_*L*_(*θ*_0_) *≠* 0, *then* ℙ*{µ*_*L*_(*θ*_0_) ∈ *I*_*µ*_(*α*)*}* → 1 − *α*.
ii. *If H*_1_(*θ*_0_), …, *H*_*L*_(*θ*_0_) *are distinct, then* ℙ*{H*_*s*_(*θ*_0_) ∈ *I*_eCI_(*α*)*}* → 1−*α, where the probability is taken over the observed data and s*.

See Section S3.3 of the Supplementary Material for the proof. Theorem 3.2(ii) holds for every *L*, and it implies that *I*_eCI_(*α*) is an asymptotic prediction interval for *H*_*s*_(*θ*_0_).

### 3.2 Mean over a population of general circulation models

We next consider the GCMs sampled from a population as in Section 2.2. For *I*_eCI_(*α*) and *I*_*µ*_(*α*), we have the following proposition on the coverage for *µ*_∞_(*θ*_0_).

#### Proposition 3.3.1

*Suppose that Assumption 1 is satisfied, that H*_*G*_ *is continuous at θ*_0_ *for every GCM G in the population, and that H*_*G*_(*θ*_0_) *follows N*(*µ*_∞_(*θ*_0_), *τ* ^2^) *with τ* ^2^ *>* 0. *Then, as n* → ∞, *the following two statements hold*.

i. *If L <* 2*/α, then* ℙ*{µ*_∞_(*θ*_0_) ∈ *I*_eCI_(*α*)*}* → 1 − 2^1−*L*^.
ii. ℙ*{µ*_∞_(*θ*_0_) ∈ *I*_*µ*_(*α*)*}* → 0.

See Section S3.4 of the Supplementary Material for the proof. The limit 1 − 2^1−*L*^ in Proposition 3.3(i) is the probability that *µ*_∞_(*θ*_0_) lies between the smallest and the largest of 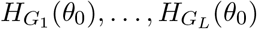. It is 0.75 for *L* = 3 and 0.9375 for *L* = 5, and it is smaller than 0.95 for *L* ≤ 5. Proposition 3.3(ii) is a consequence of the construction of *I*_*µ*_(*α*) as an interval aligned with *µ*_*L*_(*θ*_0_), which does not account for the sampling of the GCMs.

In contrast, for the intervals aligned with *µ*_∞_(*θ*_0_) and *H*_new_(*θ*_0_), we have the following theorem.

#### Theorem 3.4.

*Suppose that the assumptions of Proposition 3.3 are satisfied, that L* ≥ 2, *and that* 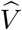 *converges to* 0 *in probability. Then, as n* → ∞, *the following two statements hold*.

i. ℙ*{µ*_∞_(*θ*_0_) ∈ *I*_RE,*t*_(*α*)*}* → 1 − *α*.
ii. ℙ*{H*_new_(*θ*_0_) ∈ *I*_new_(*α*)*}* → 1 − *α*.

See Section S3.5 of the Supplementary Material for the proof.

### 3.3 Exact coverage in finite samples under the location-shift model

In Sections 3.1 and 3.2, we considered the limits as the sample size increases, in which the differences among *H*_1_(*θ*_0_), …, *H*_*L*_(*θ*_0_) are large relative to the standard errors of 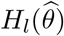. To examine the consequences of the construction of the intervals when these differences are comparable to the standard errors, we consider the location-shift model

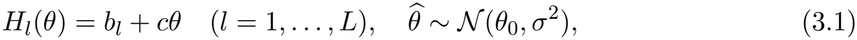

where *p* = 1, *b*_*l*_ ∈ ℝ and *c >* 0 are constants, *σ >* 0 is known, and 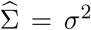. In this model, the differences among the GCM-specific impacts do not depend on *θ*, and the coverage can be computed exactly. We examine the case in which these differences depend on the coefficients in the simulation study in Section 4. Under model (3.1), the coverage of *I*_eCI_(*α*) for *µ*_*L*_(*θ*_0_) depends on the GCMs only through *e*_*l*_ = *{H*_*l*_(*θ*_0_) − *µ*_*L*_(*θ*_0_)*}/*(*cσ*) (*l* = 1, …, *L*). See Section S4.1 of the Supplementary Material for the expression of the coverage.

Figure 1(a) and (b) show the coverage of *I*_eCI_(*α*) for *µ*_*L*_(*θ*_0_) with one GCM distant from the others, i.e., *b*_1_ = *· · ·* = *b*_*L*−1_ = 0 and *b*_*L*_ = *cσβ*. The coverage was at least 1 − *α* for every value of *β* if *L* ≤ 2*/α*. If *L >* 2*/α*, it was above 1 − *α* for small values of *β* but decreased for large values of *β*, e.g., to below 0.001 for *L* ∈ *{*41, 50*}* at 1 − *α* = 0.95 and *β* = 300. See Section S4.2 of the Supplementary Material for the coverage in other configurations.

**Figure 1.**
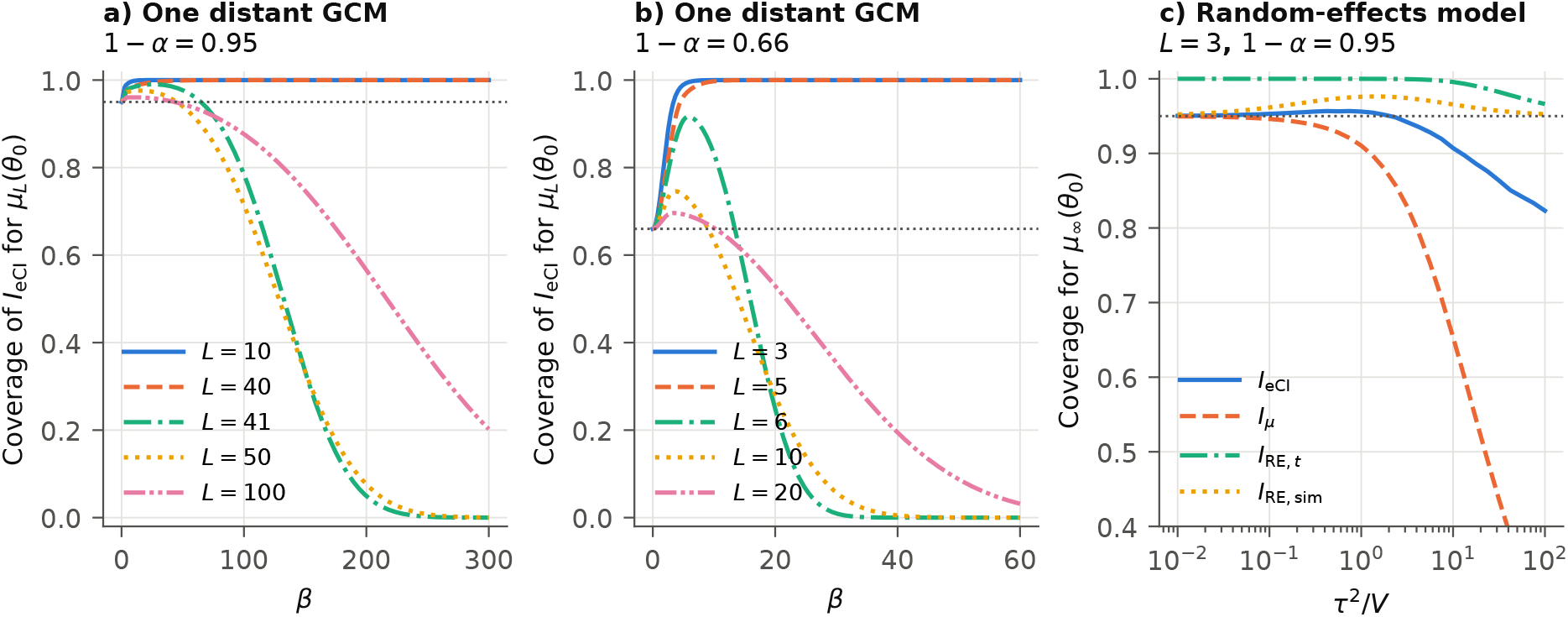
Exact coverage under model (3.1). (a, b) Coverage of *I*_eCI_(*α*) for *µ*_*L*_(*θ*_0_) with *b*_1_ = *· · ·* = *b*_*L*−1_ = 0 and *b*_*L*_ = *cσβ* for 1 − *α* = 0.95 and 0.66. (c) Coverage of the intervals at 1 − *α* = 0.95 for *µ*_∞_(*θ*_0_) with *L* = 3 under the normal random-effects model as functions of *τ* ^2^*/V* . The dotted lines are at the nominal levels.

For *µ*_∞_(*θ*_0_), we consider the normal random-effects model, in which *b*_1_, …, *b*_*L*_ in model (3.1) are independently distributed as *N*(*b*_0_, *τ* ^2^), and *V* = *c*^2^*σ*^2^. In Figure 1(c) with *L* = 3, the coverage of *I*_*µ*_(0.05) decreased as *τ* ^2^*/V* increased, that of *I*_eCI_(0.05) was below 0.95 for *τ* ^2^*/V* ≥ 2.37 and approximately 0.82 at *τ* ^2^*/V* = 100, and that of *I*_RE,*t*_(0.05) was at least 0.966. See Section S4.3 of the Supplementary Material for the expressions of the coverage and the results with *L* ∈ *{*5, 10, 20*}*.

## 4 Simulation study

In the simulation study, we compared the coverage of the intervals in a setting that mimics projections of heat-related mortality, in which the impact is a nonlinear function of the coefficients, the differences among the GCM-specific impacts depend on the coefficients, and the coefficients are estimated by the quasi-likelihood method. We also compared the intervals for the mean over a population of GCMs and for the impact under a new GCM.

### 4.1 Data generation

We generated the daily mean temperature *T*_*i*_ (*i* = 1, …, 3650) over 10 years by

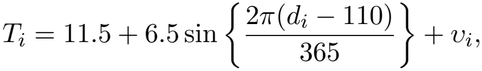

where *d*_*i*_ = (*i* − 1) mod 365, and *υ*_1_, *υ*_2_, … is the stationary first-order autoregressive process with the coefficient 0.75 and the marginal standard deviation 2.6, i.e., *υ*_1_ ∼ *N*(0, 2.6^2^) and *υ*_*i*_ − 0.75*υ*_*i*−1_ (*i* ≥ 2) are independently distributed as *N*(0, 2.6^2^(1 − 0.75^2^)). Let *f*_*θ*_(*T*) = *{B*(*T*) − *B*(18)*}*^⊤^*θ* denote the log relative risk at the temperature *T* relative to the reference temperature 18, where *B*(*T*) ∈ ℝ^4^ is the natural cubic spline basis with the knots at the 10th, 75th, and 90th percentiles and the boundary knots at the 0.1th and 99.9th percentiles of the temperature distribution. We generated the daily number of deaths *Y*_*i*_ independently given *T*_*i*_ and *ξ*_*i*_ by

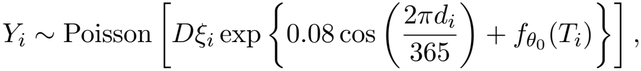

where *D* ∈ *{*20, 150*}* is the expected number of daily deaths at the reference temperature, and *ξ*_1_, *ξ*_2_, … are independently distributed as the gamma distribution with the shape 50 and the mean 1. We set the true coefficient vector *θ*_0_ so that the log relative risk 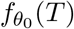 is close to 0.0009(18 − *T*)^2^ for *T <* 18 and 0.0045(*T* − 18)^2^ for *T* ≥ 18, i.e., *θ*_0_ is the value of *θ* with the smallest sum of the squared differences between *f*_*θ*_(*T*) and this function at 1,000 equally spaced values of *T* between the boundary knots. See Section S5 of the Supplementary Material for the knots and the computation of the percentiles.

For each GCM *l*, we generated the daily temperature over a future period of 21 years by

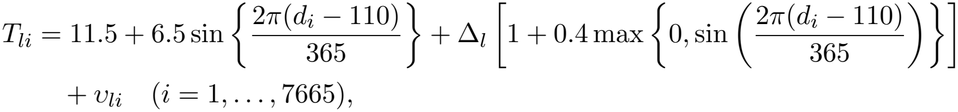

where Δ_*l*_ is the warming of GCM *l*, which is amplified in summer, and *υ*_*l*1_, *υ*_*l*2_, … is generated independently in the same manner as *υ*_1_, *υ*_2_, We defined the impact as the annual number of heat-attributable deaths

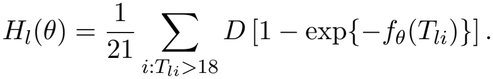

For the ensembles with the fixed GCMs, which we refer to as the fixed ensembles, we considered the following three designs, in which the GCM-specific temperature series were fixed across the repetitions. In the spread design, Δ_*l*_ = 2.5 + 0.8*z*_(*l*−0.5)*/L*_ (*l* = 1, …, *L*) with *L* ∈ *{*3, 5, 10, 20, 50*}*, where *z*_*u*_ denotes the *u*-quantile of the standard normal distribution. In the hot-model design, Δ_*l*_ = 2.5 + 0.4*z*_(*l*−0.5)*/*(*L*−1)_ (*l* = 1, …, *L* − 1) and Δ_*L*_ = 6 with *L* ∈ *{*5, 10, 20, 50*}*, which represents an ensemble including a GCM with a high equilibrium climate sensitivity (Hausfather et al., 2022). In the large-ensemble design, Δ_1_ = *· · ·* = Δ_*L*−1_ = 2.5 and Δ_*L*_ = 6 with *L* ∈ *{*5, 10, 20, 50*}*, which represents an ensemble consisting of one other GCM and the members of an initial-condition large ensemble of one GCM (Deser et al., 2020). The members of the large ensemble differ only in the internal variability.

For the ensembles with the randomly sampled GCMs, which we refer to as the random ensembles, we generated a population of 1,000 GCMs with Δ_*l*_ drawn from *N*(2.5, 0.8^2^), whose temperature series were fixed across the repetitions, and in each repetition, we sampled *L* ∈ *{*3, 5, 10, 20*}* GCMs and one new GCM with replacement from the population, so that the GCMs were sampled independently as in Section 3.2. The estimand *µ*_∞_(*θ*_0_) is the mean of *H*_*l*_(*θ*_0_) over the population.

### 4.2 Methods

We estimated *θ* by the quasi-likelihood method with the variance proportional to the mean and the mean model

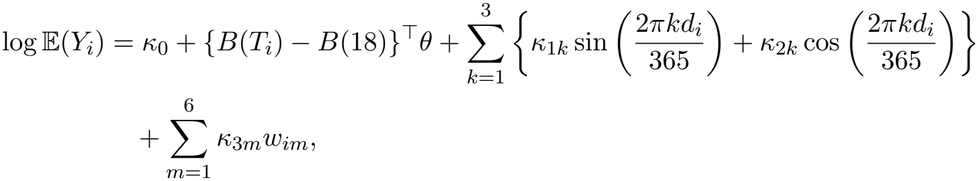

where *κ*_0_, *κ*_1*k*_, *κ*_2*k*_ (*k* = 1, 2, 3), and *κ*_3*m*_ (*m* = 1, …, 6) are the nuisance parameters, and *w*_*im*_ is 1 if (*i* − 1) mod 7 = *m* and 0 otherwise, which indicates the day of the week. This mean model has the components of the models in time series studies of temperature and mortality (Bhaskaran et al., 2013), with the seasonality represented by the harmonic terms and without the lagged temperatures, in accordance with the data generation. We estimated the covariance matrix of 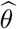 by the inverse of the negative Hessian matrix of the Poisson log-likelihood function multiplied by the dispersion parameter estimated by the Pearson statistic, as in the tutorials of Vicedo-Cabrera et al. (2019) and Quijal-Zamorano et al. (2026). We drew *J* = 1,000 coefficient vectors from 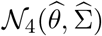 and computed the intervals at 1 − *α* ∈ *{*0.95, 0.90, 0.66*}*, for which *L >* 2*/α* holds for *L* = 50 at all levels and for *L* ≥ 10 at 1 − *α* = 0.66. We computed the empirical quantiles by the linear interpolation of type 7 of Hyndman and Fan (1996), which is the default in R and NumPy in Python.

For the fixed ensembles, we evaluated the coverage of *I*_eCI_(*α*) and *I*_*µ*_(*α*) for *µ*_*L*_(*θ*_0_), the ratio of the median width of *I*_eCI_(*α*) to that of *I*_*µ*_(*α*), and the coverage of *I*_eCI_(*α*) for *H*_*s*_(*θ*_0_). For the random ensembles, we evaluated the coverage of *I*_eCI_(*α*), *I*_*µ*_(*α*), *I*_RE,*t*_(*α*), and *I*_RE,sim_(*α*) for *µ*_∞_(*θ*_0_) and the coverage of *I*_eCI_(*α*) and *I*_new_(*α*) for *H*_new_(*θ*_0_). We repeated the data generation 2,000 times for each setting.

### 4.3 Results

Table 2 shows the coverage for the fixed ensembles with *D* = 150. See Section S5 of the Supplementary Material for the results with *D* = 20 and the coverage for *H*_*s*_(*θ*_0_). The coverage of *I*_*µ*_(*α*) for *µ*_*L*_(*θ*_0_) was close to the nominal level in all designs, between 0.960 and 0.962 at 1 − *α* = 0.95 and between 0.668 and 0.670 at 1 − *α* = 0.66. The coverage of *I*_eCI_(*α*) for *H*_*s*_(*θ*_0_) was close to the nominal level with *D* = 150, between 0.950 and 0.955 at 1 − *α* = 0.95 and between 0.651 and 0.675 at 1 − *α* = 0.66, whereas with *D* = 20, it was below the nominal level in some settings, e.g., 0.627 at 1 − *α* = 0.66 for *L* = 5 in the large-ensemble design.

**Table 2:**
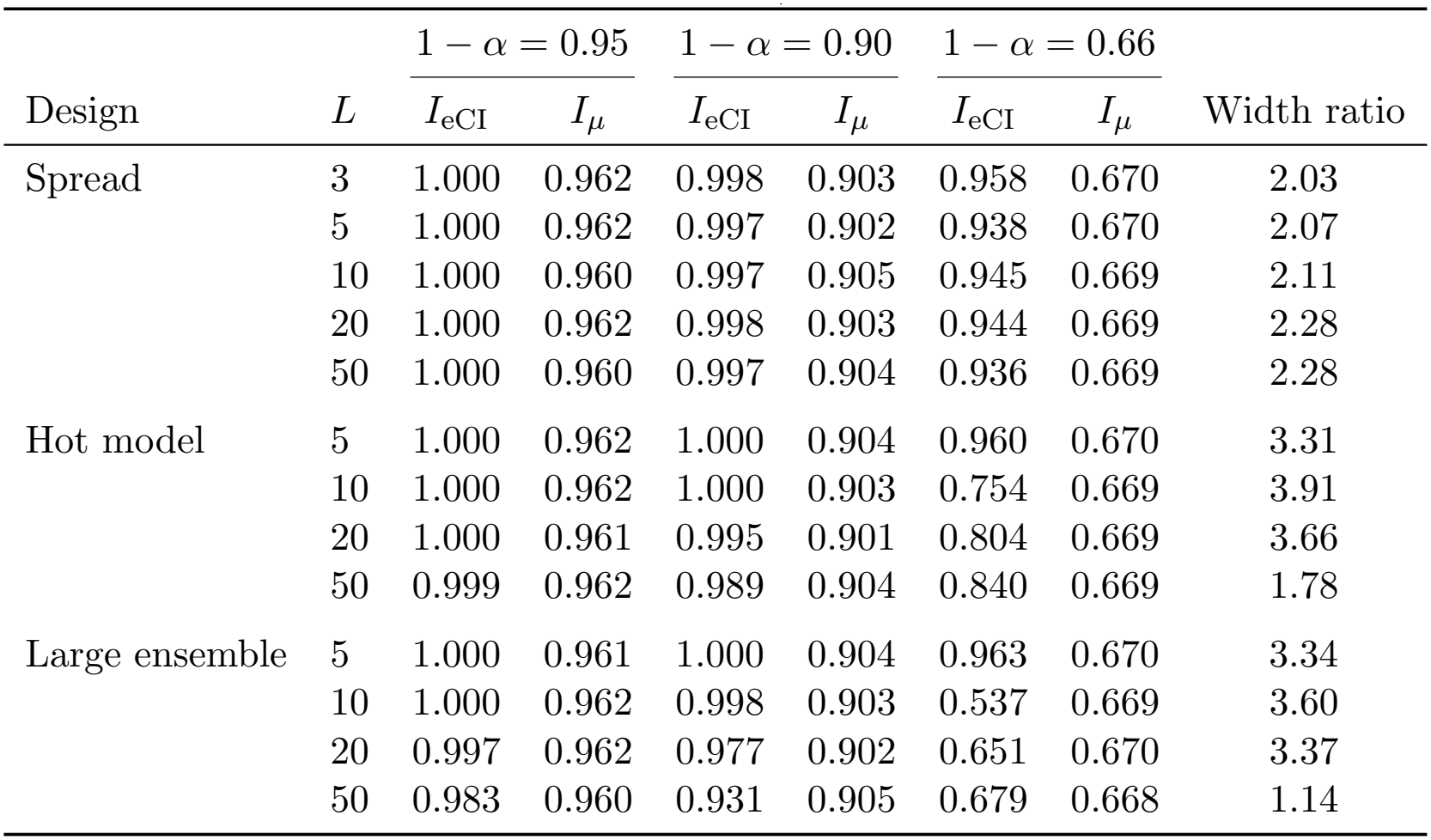
Coverage of *I*_eCI_(*α*) and *I*_*µ*_(*α*) for *µ*_*L*_(*θ*_0_) in the fixed ensembles with *D* = 150, and the ratio of the median width of *I*_eCI_(0.05) to that of *I*_*µ*_(0.05).

| Design | *L* | 1 − α = 0.95 |  | 1 − α = 0.90 |  | 1 − α = 0.66 |  |  |
|---|---|---|---|---|---|---|---|---|
|  |  | *I*<sub>eCI</sub> | *I*<sub>μ</sub> | *I*<sub>eCI</sub> | *I*<sub>μ</sub> | *I*<sub>eCI</sub> | *I*<sub>μ</sub> | Width ratio |
| Spread | 3 | 1.000 | 0.962 | 0.998 | 0.903 | 0.958 | 0.670 | 2.03 |
|  | 5 | 1.000 | 0.962 | 0.997 | 0.902 | 0.938 | 0.670 | 2.07 |
|  | 10 | 1.000 | 0.960 | 0.997 | 0.905 | 0.945 | 0.669 | 2.11 |
|  | 20 | 1.000 | 0.962 | 0.998 | 0.903 | 0.944 | 0.669 | 2.28 |
|  | 50 | 1.000 | 0.960 | 0.997 | 0.904 | 0.936 | 0.669 | 2.28 |
| Hot model | 5 | 1.000 | 0.962 | 1.000 | 0.904 | 0.960 | 0.670 | 3.31 |
|  | 10 | 1.000 | 0.962 | 1.000 | 0.903 | 0.754 | 0.669 | 3.91 |
|  | 20 | 1.000 | 0.961 | 0.995 | 0.901 | 0.804 | 0.669 | 3.66 |
|  | 50 | 0.999 | 0.962 | 0.989 | 0.904 | 0.840 | 0.669 | 1.78 |
| Large ensemble | 5 | 1.000 | 0.961 | 1.000 | 0.904 | 0.963 | 0.670 | 3.34 |
|  | 10 | 1.000 | 0.962 | 0.998 | 0.903 | 0.537 | 0.669 | 3.60 |
|  | 20 | 0.997 | 0.962 | 0.977 | 0.902 | 0.651 | 0.670 | 3.37 |
|  | 50 | 0.983 | 0.960 | 0.931 | 0.905 | 0.679 | 0.668 | 1.14 |

At 1 − *α* = 0.95, the coverage of *I*_eCI_(0.05) for *µ*_*L*_(*θ*_0_) was close to one rather than the nominal level. Over the 26 settings with *D* ∈ *{*20, 150*}*, it was 1.000 in 10 settings and at least 0.99 in 22 settings, and with *D* = 150, the median width of *I*_eCI_(0.05) was 1.14 to 3.91 times that of *I*_*µ*_(0.05). This was also the case for *L* = 50 *>* 2*/α*, for which *µ*_*L*_(*θ*_0_) was between the second smallest and the second largest of *H*_1_(*θ*_0_), …, *H*_*L*_(*θ*_0_) in all designs, i.e., the condition in Theorem 3.1(i) was satisfied. At 1 − *α* = 0.90, the coverage of *I*_eCI_(0.10) was at least the nominal level in all settings.

At 1 − *α* = 0.66, for which 2*/α* ≈ 5.9, the coverage of *I*_eCI_(0.34) with *D* = 150 decreased for *L >* 2*/α* in the designs including the GCM with Δ_*L*_ = 6. In the large-ensemble design, it was 0.963 for *L* = 5, whereas it was 0.537, 0.651, and 0.679 for *L* = 10, 20, and 50, respectively, of which only the coverage for *L* = 10 was below the nominal level by more than the Monte Carlo standard error. In the hot-model design, it was 0.960 for *L* = 5 and between 0.754 and 0.840 for *L* ≥ 10, and in the spread design, it was at least 0.936. With *D* = 20, for which the standard errors of 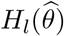 are larger relative to the differences among the GCM-specific impacts, the coverage of *I*_eCI_(0.34) was at least 0.664 in all designs.

Table 3 shows the results for the random ensembles at 1 − *α* = 0.95. See Section S5 of the Supplementary Material for the results at 1 − *α* = 0.66. For *µ*_∞_(*θ*_0_), the coverage of *I*_RE,*t*_(0.05) was at least 0.967 and that of *I*_RE,sim_(0.05) was between 0.946 and 0.953, whereas that of *I*_*µ*_(0.05) was between 0.769 and 0.938. The coverage of *I*_eCI_(0.05) was 0.947 and 0.924 for *L* = 3 with *D* = 20 and *D* = 150, respectively, and at least 0.976 for *L* ≥ 5. For *H*_new_(*θ*_0_), the coverage of *I*_eCI_(0.05) was between 0.758 and 0.935, whereas that of *I*_new_(0.05) was at least 0.947. The median width of *I*_RE,*t*_(0.05) was approximately twice that of *I*_eCI_(0.05) for *L* = 3 and between 0.53 and 0.81 times for *L* ≥ 10.

**Table 3:**
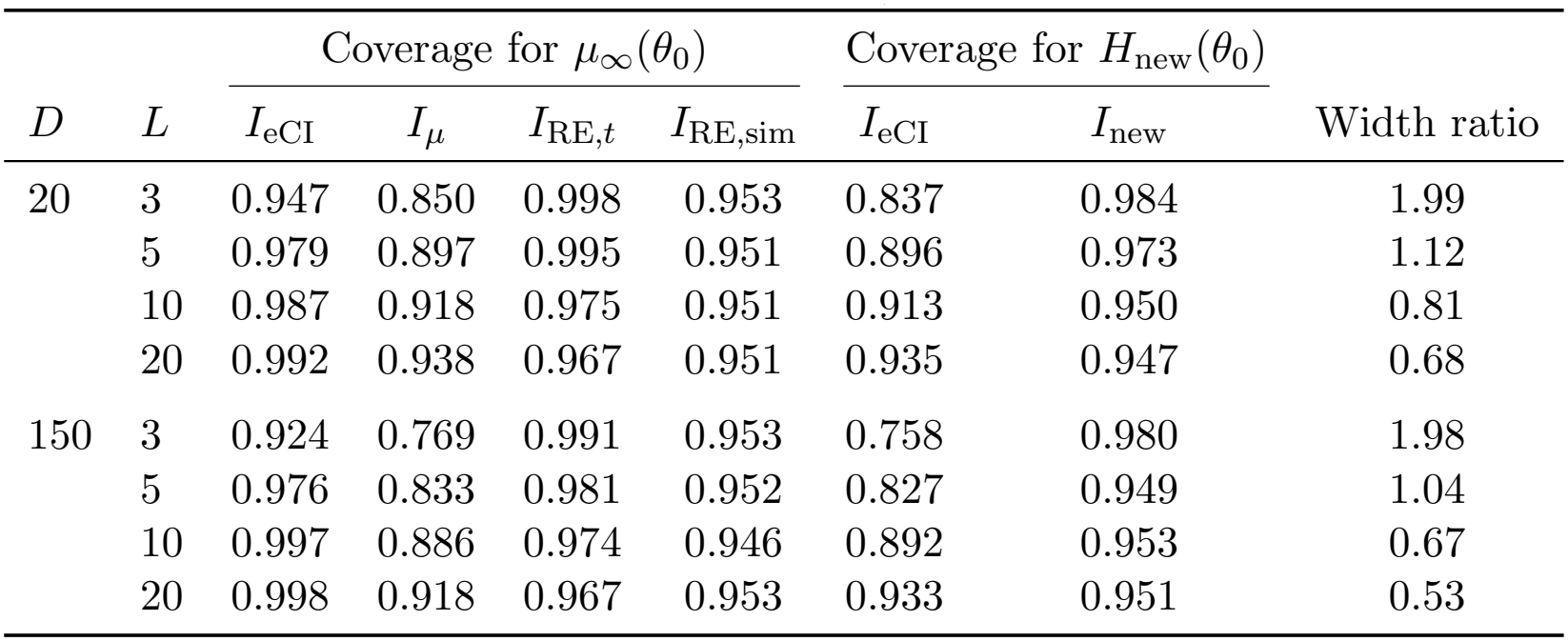
Coverage of the intervals at 1 − *α* = 0.95 for *µ*_∞_(*θ*_0_) and *H*_new_(*θ*_0_) in the random ensembles, and the ratio of the median width of *I*_RE,*t*_(0.05) to that of *I*_eCI_(0.05).

|  |  | Coverage for *μ*<sub>∞</sub>(*θ*<sub>0</sub>) |  |  |  | Coverage for *H*<sub>new</sub>(*θ*<sub>0</sub>) |  |  |
|---|---|---|---|---|---|---|---|---|
| *D* | *L* | *I*<sub>eCI</sub> | *I*<sub>*μ*</sub> | *I*<sub>RE,*t*</sub> | *I*<sub>RE,sim</sub> | *I*<sub>eCI</sub> | *I*<sub>new</sub> | Width ratio |
| 20 | 3 | 0.947 | 0.850 | 0.998 | 0.953 | 0.837 | 0.984 | 1.99 |
|  | 5 | 0.979 | 0.897 | 0.995 | 0.951 | 0.896 | 0.973 | 1.12 |
|  | 10 | 0.987 | 0.918 | 0.975 | 0.951 | 0.913 | 0.950 | 0.81 |
|  | 20 | 0.992 | 0.938 | 0.967 | 0.951 | 0.935 | 0.947 | 0.68 |
| 150 | 3 | 0.924 | 0.769 | 0.991 | 0.953 | 0.758 | 0.980 | 1.98 |
|  | 5 | 0.976 | 0.833 | 0.981 | 0.952 | 0.827 | 0.949 | 1.04 |
|  | 10 | 0.997 | 0.886 | 0.974 | 0.946 | 0.892 | 0.953 | 0.67 |
|  | 20 | 0.998 | 0.918 | 0.967 | 0.953 | 0.933 | 0.951 | 0.53 |

Note that, with the 2,000 repetitions, the Monte Carlo standard error of the coverage is approximately 0.005 at 0.95 and 0.011 at 0.66.

## 5 Reanalysis of heat-related mortality projections in London

### 5.1 Data

We reanalyzed the projections of heat-related mortality in London under SSP2-4.5 in the tutorial of Quijal-Zamorano et al. (2026). In the tutorial, the age-specific exposure–response functions for persons younger than 75 years and those aged 75 years or older were estimated by the quasi-likelihood method with the variance proportional to the mean, in which the mean of the daily number of deaths was modeled by a distributed lag nonlinear model (Gasparrini et al., 2010). The daily numbers of heat-attributable deaths from 1950 to 2099 were projected with the daily temperature trajectories of *L* = 3 GCMs and *J* = 100 coefficient draws. We used the daily attributable numbers of deaths at the point estimate and the coefficient draws for each GCM and each age group, which are available from the repository of the tutorial.

### 5.2 Methods

The target of the reported mean depends on the purpose of the projection. The mean *µ*_*L*_(*θ*_0_) over the three GCMs is the target when the purpose is the projection under these GCMs, and the mean *µ*_∞_(*θ*_0_) over a population of GCMs is the target when the three GCMs are regarded as a sample representing the differences among climate models. The eCI is the interval for *H*_*s*_(*θ*_0_), the impact under one of the three GCMs selected uniformly at random.

For the two age groups and the total population, we computed the annual heat-attributable deaths in each decade and in two 21-year periods, the GCM-specific window centered on the year in which the global warming level of 2 ^°^C is reached and the period 2079–2099. For each of them, we computed 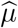, *I*_*µ*_(*α*), and *I*_RE,*t*_(*α*) at 1 − *α* ∈ *{*0.95, 0.66*}*, where the empirical quantiles were computed as in Section 4.2. The values of 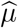 and *I*_eCI_(0.05) agreed with those reported in the tutorial. For the two 21-year periods, we estimated the Monte Carlo standard errors of the endpoints of the intervals by the standard deviations over 2,000 resamples of the *J* coefficient draws with replacement, in which the impacts under the GCMs with the same draw were resampled together. For the two 21-year periods, we also computed 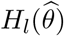 and the 2.5th and 97.5th percentiles of *H*_*l*_(*θ*^(*j*)^) over the draws for each GCM, and we decomposed the variance of the pooled impacts *H*_*l*_(*θ*^(*j*)^) (*l* = 1, …, *L*; *j* = 1, …, *J*), from which the quantiles in *I*_eCI_(*α*) are computed, into the mean of the variances over the draws within the GCMs and the variance of the means over the draws among the GCMs, which is analogous to the partition of the uncertainty in climate predictions into its sources (Hawkins and Sutton, 2009).

### 5.3 Results

Figure 2 and Table 4 show the results. At 1 − *α* = 0.95, the difference in the widths of *I*_eCI_(0.05) and *I*_*µ*_(0.05) increased in later decades, particularly for persons aged 75 years or older and the total population. For the total population, *I*_eCI_(0.05) and *I*_*µ*_(0.05) were (906, 1883) and (1060, 1520) for the 2 ^°^C warming level and (906, 3033) and (1434, 2061) for 2079–2099, respectively. For persons aged 75 years or older and the total population, the differences between the corresponding endpoints of *I*_eCI_(0.05) and *I*_*µ*_(0.05) in the two 21-year periods were at least 4.8 times the larger of the Monte Carlo standard errors of the two endpoints. For persons younger than 75 years, the differences between the lower limits were less than twice the Monte Carlo standard errors, so that the comparison of these endpoints is not reliable with *J* = 100.

**Table 4:**
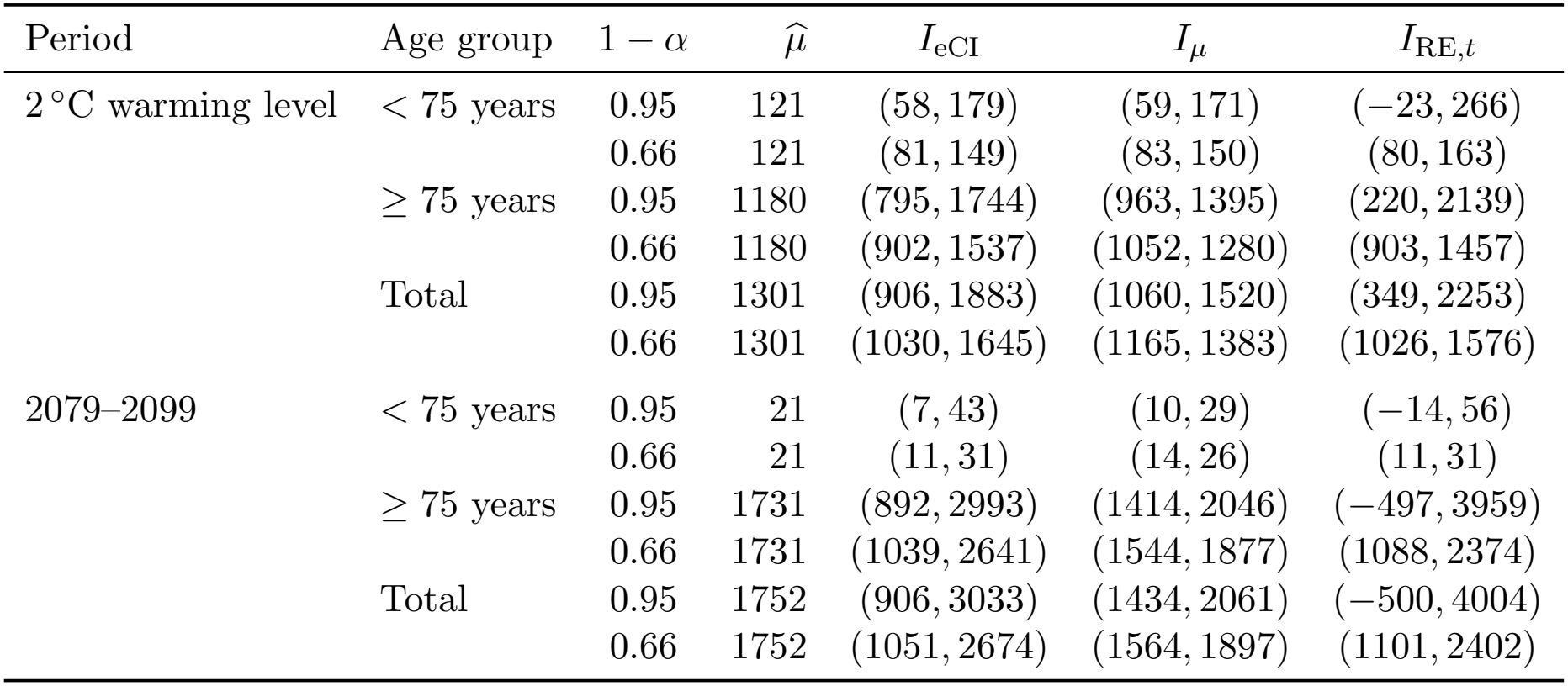
Point estimates and intervals of the annual heat-related deaths in London in the two 21-year periods.

| Period | Age group | 1 − α | μ̂ | *I*<sub>eCI</sub> | *I*<sub>μ</sub> | *I*<sub>RE,*t*</sub> |
|---|---|---|---|---|---|---|
| 2 °C warming level | < 75 years | 0.95 | 121 | (58, 179) | (59, 171) | (−23, 266) |
|  |  | 0.66 | 121 | (81, 149) | (83, 150) | (80, 163) |
|  | ≥ 75 years | 0.95 | 1180 | (795, 1744) | (963, 1395) | (220, 2139) |
|  |  | 0.66 | 1180 | (902, 1537) | (1052, 1280) | (903, 1457) |
|  | Total | 0.95 | 1301 | (906, 1883) | (1060, 1520) | (349, 2253) |
|  |  | 0.66 | 1301 | (1030, 1645) | (1165, 1383) | (1026, 1576) |
| 2079–2099 | < 75 years | 0.95 | 21 | (7, 43) | (10, 29) | (−14, 56) |
|  |  | 0.66 | 21 | (11, 31) | (14, 26) | (11, 31) |
|  | ≥ 75 years | 0.95 | 1731 | (892, 2993) | (1414, 2046) | (−497, 3959) |
|  |  | 0.66 | 1731 | (1039, 2641) | (1544, 1877) | (1088, 2374) |
|  | Total | 0.95 | 1752 | (906, 3033) | (1434, 2061) | (−500, 4004) |
|  |  | 0.66 | 1752 | (1051, 2674) | (1564, 1897) | (1101, 2402) |

**Figure 2.**
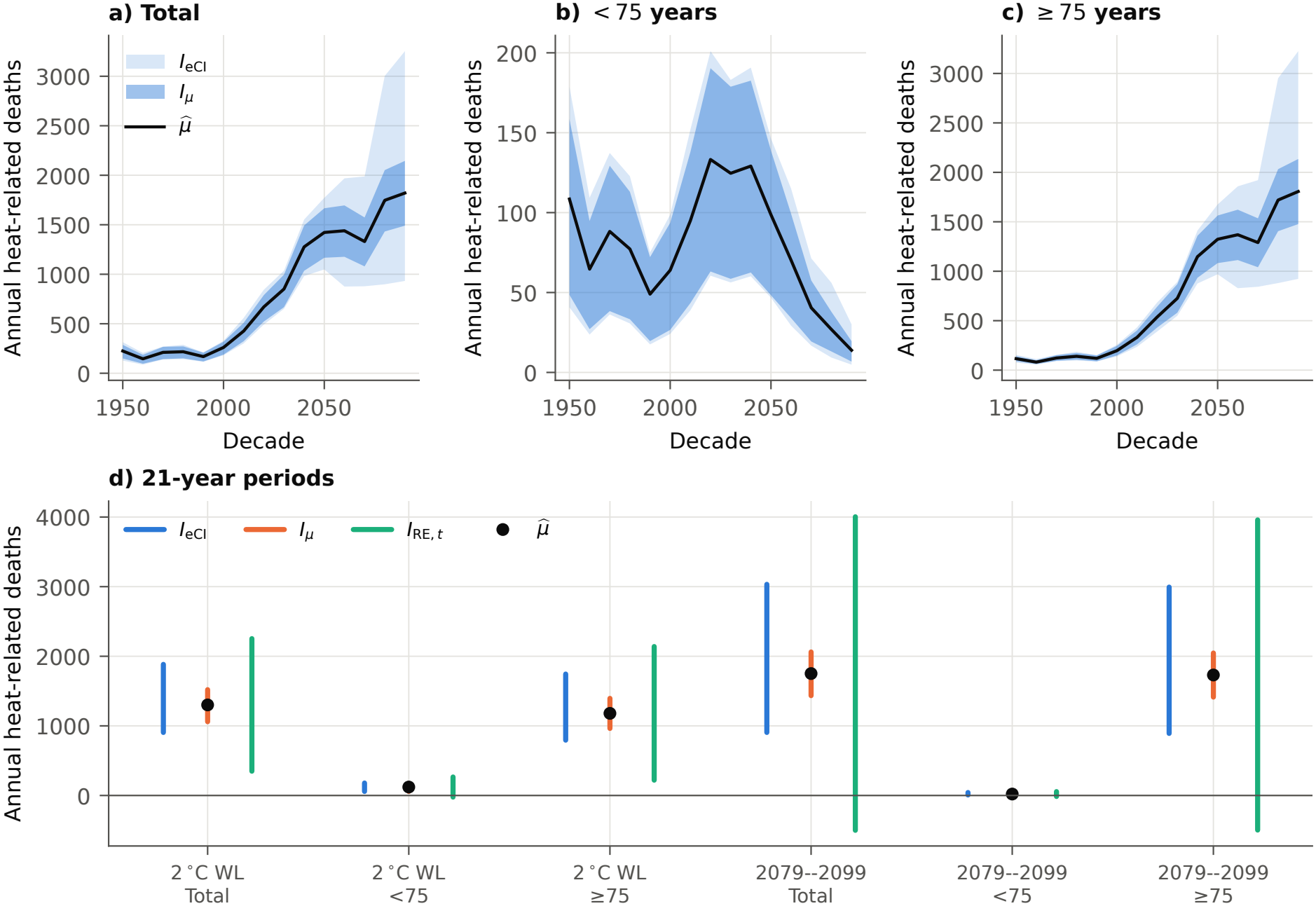
Reanalysis of the projected annual heat-related deaths in London under SSP2-4.5 with three GCMs. (a–c) Annual heat-related deaths in each decade for the total population, persons younger than 75 years, and persons aged 75 years or older. The solid lines are 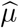, and the lighter and darker bands are *I*_eCI_(0.05) and *I*_*µ*_(0.05), respectively. (d) Annual heat-related deaths in the 21-year periods for the 2 ^°^C global warming level (WL) and 2079–2099, with *I*_eCI_(0.05), *I*_*µ*_(0.05), and *I*_RE,*t*_(0.05).

For 2079–2099, 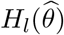 for the total population was 2714, 1478, and 1063 under ACCESS-CM2, BCC-CSM2-MR, and CESM2, respectively, and the percentile interval for ACCESS-CM2, (2247, 3175), did not overlap with those for BCC-CSM2-MR and CESM2, (1189, 1756) and (866, 1254). The variance among the GCMs accounted for 94% of the variance of the pooled impacts for 2079–2099 and 82% for the 2 ^°^C warming level. See Section S6 of the Supplementary Material for the results for each GCM and each age group. For 2079–2099, the upper and lower limits of *I*_eCI_(0.05) were determined only by the draws under ACCESS-CM2 and CESM2, respectively. Thus, the upper limit 3033 of *I*_eCI_(0.05) represents the burden under the GCM with the largest impact rather than the uncertainty in the mean over the three GCMs, for which the upper limit of *I*_*µ*_(0.05) was 2061.

Since *L* = 3 *<* 2*/α* for both levels, the coverage of *I*_eCI_(*α*) for *µ*_*L*_(*θ*_0_) converges to 1 by Theorem 3.1(i) unless the GCM-specific impacts are all equal. However, the standard deviations of *H*_*l*_(*θ*^(*j*)^) over the draws for the total population ranged from 101 to 152 for the 2 ^°^C warming level and from 104 to 247 for 2079–2099 among the three GCMs, so that the differences among the GCM-specific impacts depend on the coefficients, and the coverage for the sample size of these data is not guaranteed by the theorem.

For *µ*_∞_(*θ*_0_), *I*_RE,*t*_(0.05) was (349, 2253) and (−500, 4004) for the total population, which were wider than *I*_eCI_(0.05) because of the quantile *t*_2,0.975_ = 4.30. The negative lower limit for 2079–2099 is due to the normal distribution of the GCM-specific impacts assumed in *I*_RE,*t*_(*α*) with *L* = 3. In the tutorial, the attributable fraction (RR − 1)*/*RR on the days above the minimum mortality temperature was not truncated at 0, where RR is the relative risk, so that the nonnegativity of *µ*_∞_(*θ*_0_) does not follow from the definition of the impacts. At 1 − *α* = 0.66, *I*_RE,*t*_(0.34) was (1026, 1576) and (1101, 2402), which were narrower than *I*_eCI_(0.34), (1030, 1645) and (1051, 2674). For the total population, the Monte Carlo standard errors of the endpoints of *I*_eCI_(*α*), *I*_*µ*_(*α*), and *I*_RE,*t*_(*α*) were at most 62 at both levels.

## 6 Discussion

In this study, we specified the targets for which the GCM-ensemble mean is reported and the intervals aligned with them. The eCI used in previous studies is aligned with the impact under a GCM selected uniformly at random from the considered set. Its nominal level does not indicate its coverage for the mean over the considered GCMs or for the mean over a population of GCMs. Table S1 of the Supplementary Material summarizes the coverage of the intervals for their targets and that of the eCI for the other targets, obtained in Sections 3 and 4.

The appropriate target depends on the purpose of each study. For example, when the purpose is the projection under the considered GCMs, the target is *µ*_*L*_(*θ*_0_), and *I*_*µ*_(*α*) is the interval aligned with this target. Since *I*_*µ*_(*α*) does not represent the differences among the GCMs, these differences can be shown separately by the GCM-specific impacts, e.g., *H*_*l*_(*θ*) with their percentile intervals as in Table S6 of the Supplementary Material. The eCI can also be useful to represent these differences, but its nominal level does not indicate its coverage probability for *µ*_*L*_(*θ*_0_), so that it should be interpreted separately from *I*_*µ*_(*α*), the asymptotic confidence interval for *µ*_*L*_(*θ*_0_). Note that, since *I*_*µ*_(*α*) is computed from the same coefficient draws as the eCI, it can be obtained in existing projections without additional computation.

The targets can also be related to the impact under the actual climate, depending on how the GCMs are related to the actual climate. Annan and Hargreaves (2010) contrasted two paradigms for interpreting multi-model ensembles in climate science. In the truth-centred paradigm, the GCMs are regarded as sampled from a distribution centered on the actual climate. If this centering also holds for the impacts, the impact under the actual climate corresponds to *µ*_∞_(*θ*_0_). In the paradigm of a statistically indistinguishable ensemble, the actual climate is regarded as exchangeable with the GCMs. If the GCMs and the actual climate are regarded as independent draws from the same population, the impact under the actual climate corresponds to *H*_new_(*θ*_0_). If the actual climate is regarded as one of the considered GCMs, each with the probability 1*/L*, the impact under the actual climate corresponds to *H*_*s*_(*θ*_0_). In practice, however, the GCMs in multi-model ensembles share components and code, and some GCMs are near-replicates of others (Knutti et al., 2013; Abramowitz et al., 2019), so that the effective number of independent GCMs may be smaller than *L*, and the population of GCMs over which *µ*_∞_(*θ*_0_) is defined may not be specified in practice (Tebaldi and Knutti, 2007).

To our knowledge, the terminology “empirical confidence interval” in this literature dates at least to Gasparrini and Leone (2014). In their setting, the coefficients were drawn from an approximate sampling distribution and mapped through the same attributable-risk functional as the point estimate, and the empirical percentiles can be interpreted as a simulation-based approximate confidence interval for the attributable risk. In the GCM ensemble, the pooling across the GCMs changes the functional through which the coefficient draws are mapped, and the interpretation of the empirical percentiles changes accordingly.

In large samples with the GCMs fixed, whether the eCI covers *µ*_*L*_(*θ*_0_) depends on *L* relative to 2*/α*. At 1 − *α* = 0.95, *L <* 40 holds in the projection studies that reported the eCI with 5 (Guo et al., 2018), 18 (Chen et al., 2024), 27 (Zhan et al., 2023), and 28 (Yang et al., 2021) GCMs, for which the eCI is conservative for the mean over the considered GCMs unless the GCM-specific impacts are all equal, and its coverage was close to one in the simulation study. In contrast, Weinberger et al. (2017) pooled 41 and 42 simulation runs of 20 GCMs under the two emission scenarios, including 10 runs of one GCM, for which *L >* 2*/α* at 1 − *α* = 0.95, and the coverage converges to 0 if the impact under one GCM is so large that *µ*_*L*_(*θ*_0_) exceeds the second largest of *H*_1_(*θ*_0_), …, *H*_*L*_(*θ*_0_). For lower nominal levels, the condition *L <* 2*/α* is more restrictive, i.e., *L <* 20 for 1 − *α* = 0.90 and *L* ≤ 5 for 1 − *α* = 0.66, and the coverage was 0.537 at 1 − *α* = 0.66 for *L* = 10 in the large-ensemble design in the simulation study.

When the GCMs are regarded as a sample from a population with normally distributed impacts, the coverage of *I*_*µ*_(*α*) for the population mean converges to 0, since *I*_*µ*_(*α*) does not account for the sampling of the GCMs, and if *L <* 2*/α*, that of the eCI converges to 1 − 2^1−*L*^. In contrast, *I*_RE,*t*_(*α*) accounts for both the estimation of the coefficients and the sampling of the GCMs, although it is wide for a small number of GCMs because of the quantile of the *t* distribution with *L* − 1 degrees of freedom, as in the reanalysis with *L* = 3.

This study has the following limitations. With a small number of coefficient draws, the endpoints of the intervals are subject to Monte Carlo error, which was small relative to the differences between the intervals for the total population in the reanalysis with *J* = 100. The coverage of *I*_*µ*_(*α*) for *µ*_*L*_(*θ*_0_) relies on the consistency of 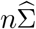 for Σ_0_ in Assumption 1, which may not hold for the estimator in Section 4.2 if the conditional variance of the daily number of deaths given the covariates is not proportional to the conditional mean. In such cases, a sandwich estimator can be used for 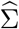 without changing the construction of the intervals. The intervals for *µ*_∞_(*θ*_0_) assume that the GCMs are sampled independently from a population. We did not consider other sources of uncertainty, such as the bias correction of the GCM outputs, the internal variability of the climate, and the demographic scenarios.

## Data Availability

The code for reproducing the numerical results is available at https://github.com/t-yui/coverage_eCI. The data for the reanalysis are available from the repository of the tutorial of Quijal-Zamorano et al. (2026) (https://github.com/marcosqz/climate_demographic_health_projections) and its fork (https://github.com/t-yui/climate_demographic_health_projections/tree/reanalysis).

https://github.com/t-yui/climate_demographic_health_projections/tree/reanalysis

## Data and code availability

The code for reproducing the numerical results is available at *https://github.com/t-yui/coverage_eCI*. The data for the reanalysis are available from the repository of the tutorial of Quijal-Zamorano et al. (2026) (*https://github.com/marcosqz/climate_demographic_health_projections*) and its fork (*https://github.com/t-yui/climate_demographic_health_projections/tree/reanalysis*).

## Ethics approval

Ethics approval was not required because this study consisted of simulation experiments and a reanalysis of publicly available, non-individual-level data.

## Supplementary Material for “Coverage of Empirical Confidence Intervals for the Ensemble Mean of General Circulation Models in Projecting Climate Change Impacts on Health”

### S1 Summary of the coverage results

Table S1 summarizes the coverage of the intervals in Table 1 of the main text for their targets and that of *I*_eCI_(*α*) for the other targets.

**Table S1:**
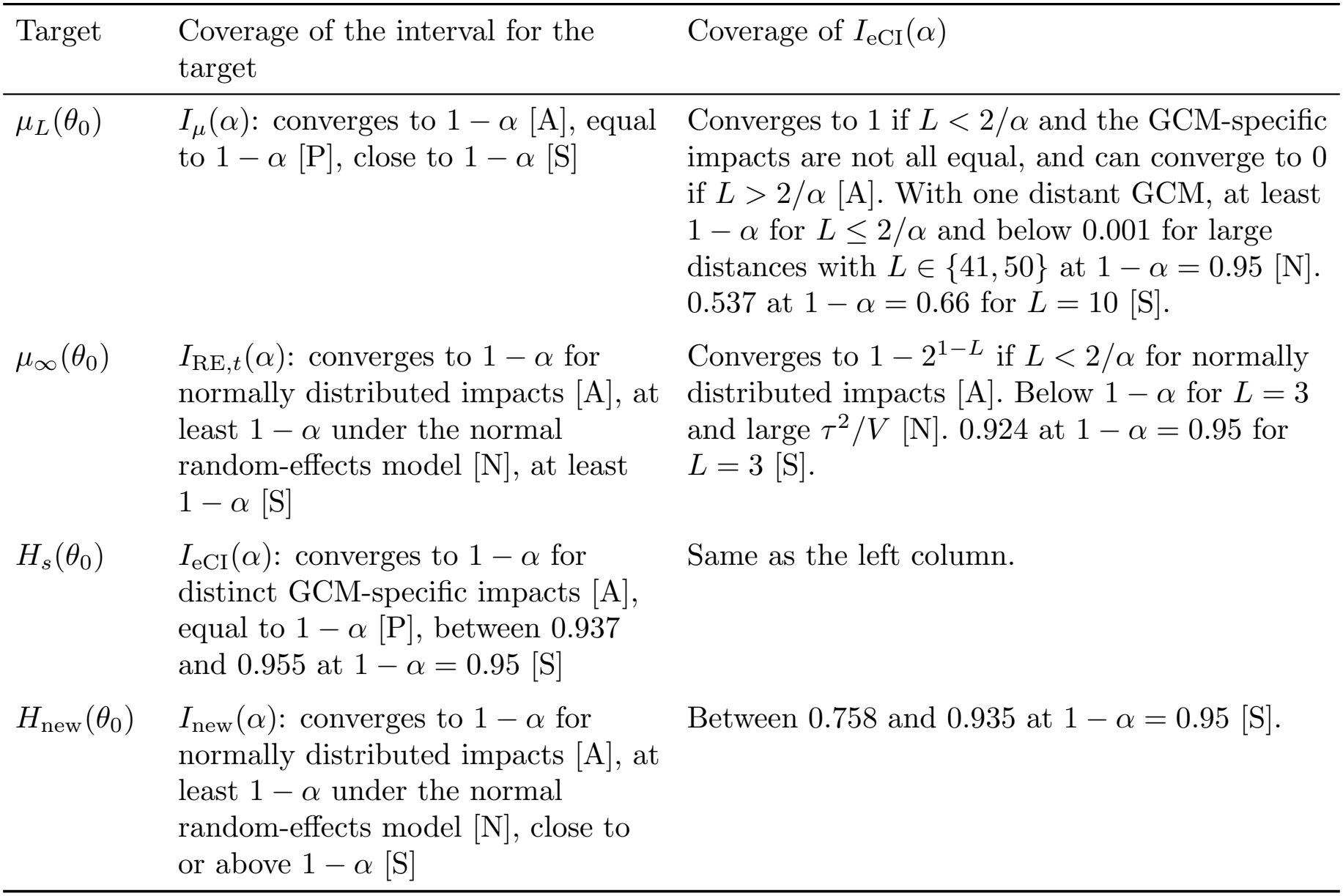
Coverage of the intervals aligned with the targets and that of *I*_eCI_(*α*). The labels indicate that the statement is proved in large samples under the regularity conditions in Section S3 without a parametric form of the maps [A], proved under model (3.1) of the main text [P], obtained by numerical computation under model (3.1) [N], or observed in the simulation study in Section 4 of the main text [S]. The statements labeled [A], [P], and [N] are for the intervals with *J* → ∞, and those labeled [S] are for *J* = 1,000.

| Target | Coverage of the interval for the target | Coverage of I<sub>eCI</sub>(α) |
|---|---|---|
| μ<sub>L</sub>(θ<sub>0</sub>) | I<sub>μ</sub>(α): converges to 1 − α [A], equal to 1 − α [P], close to 1 − α [S] | Converges to 1 if L < 2/α and the GCM-specific impacts are not all equal, and can converge to 0 if L > 2/α [A]. With one distant GCM, at least 1 − α for L ≤ 2/α and below 0.001 for large distances with L ∈ {41, 50} at 1 − α = 0.95 [N]. 0.537 at 1 − α = 0.66 for L = 10 [S]. |
| μ<sub>∞</sub>(θ<sub>0</sub>) | I<sub>RE,t</sub>(α): converges to 1 − α for normally distributed impacts [A], at least 1 − α under the normal random-effects model [N], at least 1 − α [S] | Converges to 1 − 2<sup>1−L</sup> if L < 2/α for normally distributed impacts [A]. Below 1 − α for L = 3 and large τ<sup>2</sup>/V [N]. 0.924 at 1 − α = 0.95 for L = 3 [S]. |
| H<sub>s</sub>(θ<sub>0</sub>) | I<sub>eCI</sub>(α): converges to 1 − α for distinct GCM-specific impacts [A], equal to 1 − α [P], between 0.937 and 0.955 at 1 − α = 0.95 [S] | Same as the left column. |
| H<sub>new</sub>(θ<sub>0</sub>) | I<sub>new</sub>(α): converges to 1 − α for normally distributed impacts [A], at least 1 − α under the normal random-effects model [N], close to or above 1 − α [S] | Between 0.758 and 0.935 at 1 − α = 0.95 [S]. |

### S2 Target distribution of the empirical confidence interval

Let 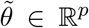 denote a random vector following 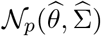, and let *s* be a random variable uniformly distributed on *{*1, …, *L}* and independent of 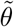. Thus, *s* is an index that assigns probability 1*/L* to each GCM. Let ℙ^*^ denote the probability under the joint distribution of 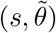 given the data. For *x* ∈ ℝ, the cumulative distribution function of 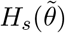 is given by

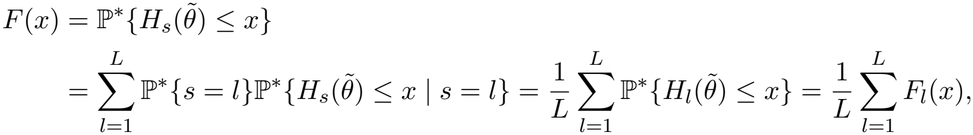

where the third equality follows from the independence of *s* and 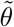, and

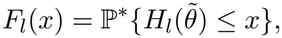

for *l* = 1, …, *L*.

We show that, as *J* increases, the empirical distribution of *ℋ* in (2.1), which is pooled across *l* ∈ *{*1, …, *L}* and *j* ∈ *{*1, …, *J}*, converges to *F* . For the coefficient vectors *θ*^(1)^, …, *θ*^(*J*)^ drawn independently from 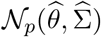, the empirical cumulative distribution function of the pooled impact values is given by

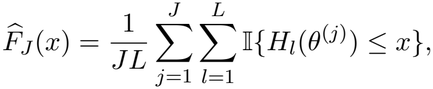

where I*{·}* is the indicator function. Define

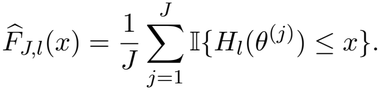

Then, since, for fixed *l, H*_*l*_(*θ*^(1)^), …, *H*_*l*_(*θ*^(*J*)^) are independent and identically distributed with the distribution function *F*_*l*_ under ℙ^*^, by the Glivenko–Cantelli theorem, as *J* → ∞, we have

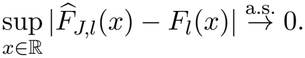

Using the triangle inequality, for *x* ∈ ℝ, we obtain

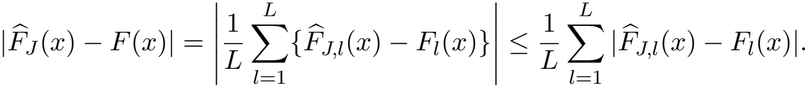

Therefore, we have

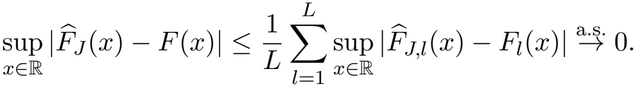

This result shows that the pooled empirical distribution converges to the equally weighted mixture distribution with the cumulative distribution function *F* .

Next, we prove the convergence of the quantiles of the empirical distribution. For 0 *< u <* 1, define

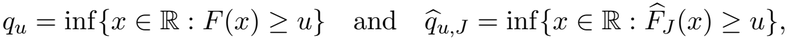

and suppose that, for every *ε >* 0,

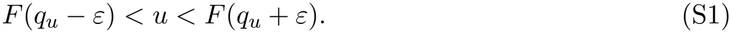

Let *ε >* 0. Since *u* − *F* (*q*_*u*_ − *ε*) *>* 0 and *F* (*q*_*u*_ + *ε*) − *u >* 0, almost surely, we have

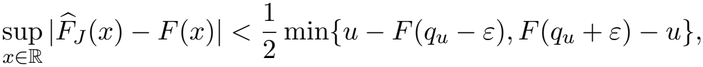

for sufficiently large *J*. Thus, we obtain

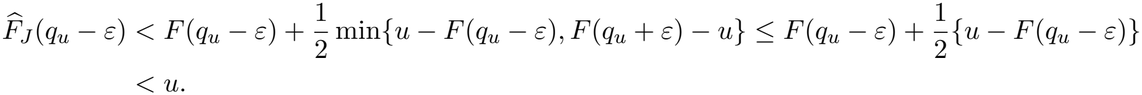

For *x* ≤ *qu* − *ε* we have 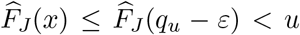. By the definition of 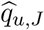, we then obtain 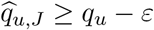. Similarly, we obtain

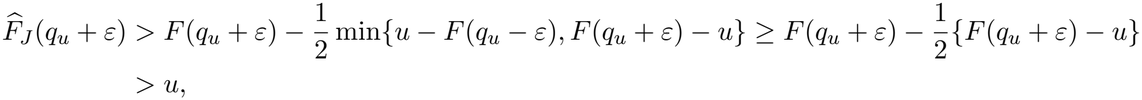

and hence 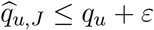. We therefore obtain

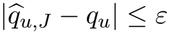

for sufficiently large *J*, almost surely, and since *ε >* 0 is arbitrary, this means

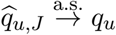

as *J* → ∞ under ℙ^*^. Consequently, provided that the *α/*2- and (1 − *α/*2)-quantiles of *F* are unique in the sense that (S1) holds for *u* = *α/*2 and *u* = 1−*α/*2, the endpoints of *I*_eCI_(*α*) converge almost surely to the corresponding quantiles of the equally weighted mixture distribution *F* . The same argument with *L* = 1 and *H*_1_ replaced by *µ*_*L*_ applies to *I*_*µ*_(*α*).

Note that, since the empirical *u*-quantile of *ℋ* with linear interpolation, which is the default in *R* and in *NumPy* for *Python*, lies between 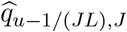 and 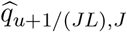, the same argument with *u* replaced by *u* − 1*/*(*JL*) and *u* + 1*/*(*JL*) shows that it also converges almost surely to *q*_*u*_.

### S3 Large-sample results

In this section, we prove Theorems 3.1, 3.2, and 3.4 and Proposition 3.3 of the main text.

#### S3.1 Notation and lemmas

Let ℙ^*^, *F*_*l*_, *F*, and *q*_*u*_ be as in Section S2. As defined in Section 2.4 of the main text, we have *I*_eCI_(*α*) = [*q*_*α/*2_, *q*_1−*α/*2_]. Let *Z*^*^ follow *N*_*p*_(0, *I*_*p*_) independently of the data, so that 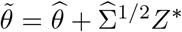. Let ℙ denote the probability under the joint distribution of the data, *Z*^*^, and *s*. Let Φ and *ϕ* denote the cumulative distribution function and the density function of *N*(0, 1). For *y* ∈ ℝ, by the definition of *q*_*u*_ and the right continuity of *F*, we have the following four implications:

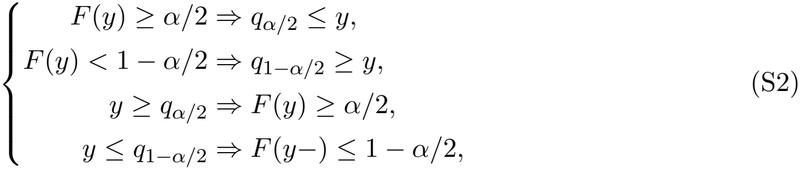

where *F* (*y*−) = lim_*x*↑*y*_ *F* (*x*).

As preparation, we prove the following two lemmas.

##### Lemma S1.

*Suppose that Assumption 1 is satisfied and that a map H* : ℝ^*p*^ → ℝ *is continuous at θ*_0_. *Then, for x > H*(*θ*_0_), 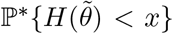 *converges to* 1 *in probability, and for x < H*(*θ*_0_), 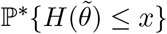 *converges to* 0 *in probability*.

*Proof*. By Assumption 1, we have 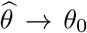 and 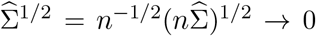 in probability, and hence 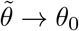 in probability under ℙ. Let *ε >* 0. Since *H* is continuous at *θ*_0_, we have

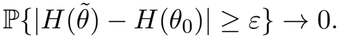

Since the expectation of 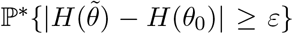 over the data is 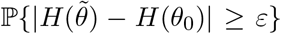, by Markov’s inequality, for *δ >* 0, we have

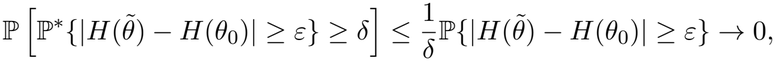

i.e.,

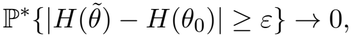

in probability. For *x > H*(*θ*_0_), with *ε* = *x* − *H*(*θ*_0_), since 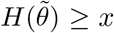 implies 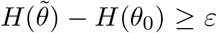, we have

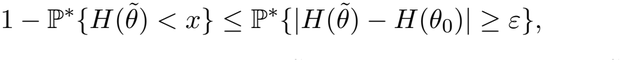

and for *x < H*(*θ*_0_), with *ε* = *H*(*θ*_0_) − *x*, since 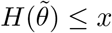 implies 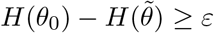, we have

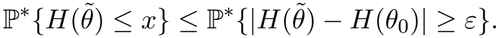

Therefore, we have the statement.

##### Lemma S2.

*Suppose that Assumption 1 is satisfied and that a map H* : ℝ^*p*^ → ℝ *is continuously differentiable in a neighborhood of θ*_0_ *with* ∇*H*(*θ*_0_) *≠* 0. *Let ς* = *{*∇*H*(*θ*_0_)^⊤^Σ_0_∇*H*(*θ*_0_)*}*^1*/*2^. *Then, the following two statements hold*.

i. *We have the following convergence, which also holds with* ≤ *replaced by <*.

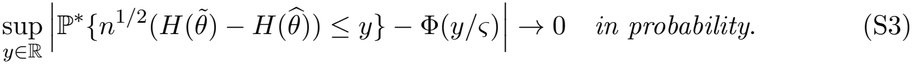

i. *The distribution of* 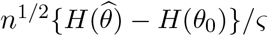 *converges to N*(0, 1).

*Proof*. Since Σ_0_ is positive definite and ∇*H*(*θ*_0_) *≠* 0, we have *ς >* 0. The statement (ii) follows from Theorem 3.1 of van der Vaart (1998), whose conditions are satisfied because *H* is differentiable at *θ*_0_ and 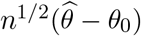 converges in distribution by Assumption 1.

Let *U* be an open ball centered at *θ*_0_ on which *H* is continuously differentiable, and let 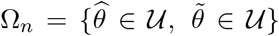. Since 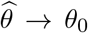 and 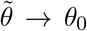 in probability under ℙ as in the proof of Lemma S1, we have ℙ(Ω_*n*_) → 1. On Ω_*n*_, by the mean value theorem, there exists 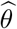 on the segment between 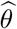 and 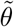 such that

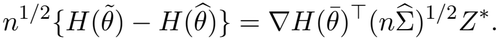

Let 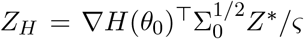, which follows *N*(0, 1) independently of the data, and let 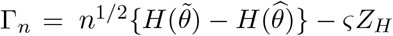. On Ω_*n*_, we have

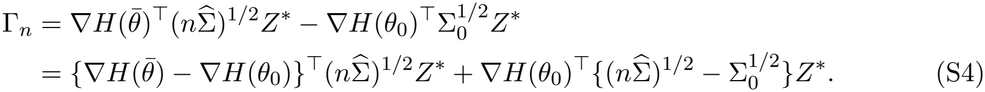

Since 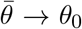 in probability, ∇*H* is continuous at *θ*_0_, and the matrix square root is continuous at Σ_0_, both terms on the right-hand side of (S4) converge to 0 in probability, and hence Γ_*n*_ → 0 in probability under ℙ. Let *ε >* 0. Since 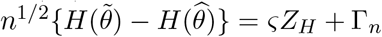, for *y* ∈ ℝ, we have

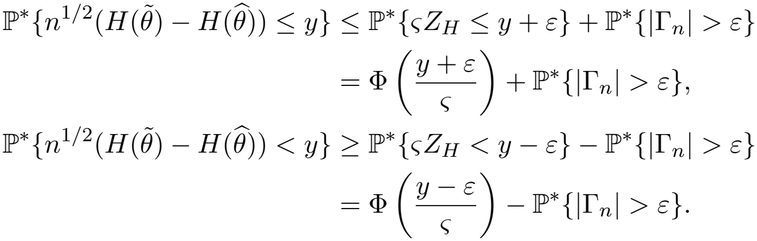

Since 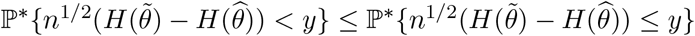 and |Φ((*y ± ε*)*/ς*) − Φ(*y/ς*)| ≤ *εϕ*(0)*/ς*, we obtain

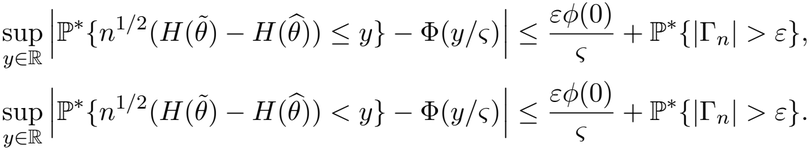

By Markov’s inequality as in the proof of Lemma S1, we have ℙ^*^*{*|Γ_*n*_| *> ε}* → 0 in probability, and since *ε >* 0 is arbitrary, we obtain (S3) and its counterpart with *<*. Thus, the statement (i) holds.

#### S3.2 Proof of Theorem 3.1

We prove Theorem 3.1 by using Lemma S1.

*Proof of Theorem 3.1*. Let *µ* = *µ*_*L*_(*θ*_0_).

We first prove (i). Let

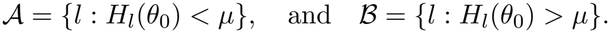

Since 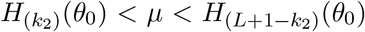, we have |*A*| ≥ *k*_2_ and |*B*| ≥ *k*_2_, where *k*_2_ *> Lα/*2. From Lemma S1, we have

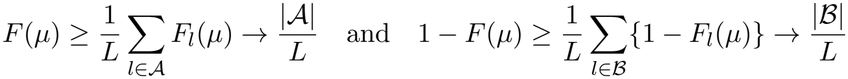

in probability. Since |*A*|*/L > α/*2 and |*B*|*/L > α/*2, we have

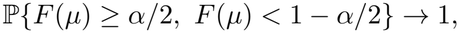

and from the first and second implications in (S2), we obtain

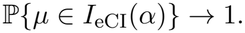

We next prove (ii). Suppose that 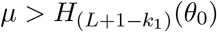, and let

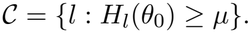

Then, at least *L* + 1 − *k*_1_ of *H*_1_(*θ*_0_), …, *H*_*L*_(*θ*_0_) are smaller than *µ*, and hence

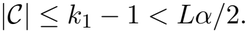

Let *x* = *{µ* + max_*l*∉C_ *H*_*l*_(*θ*_0_)*}/*2, which satisfies *H*_*l*_(*θ*_0_) *< x < µ* for *l* ∉ *C*. From Lemma S1, we have

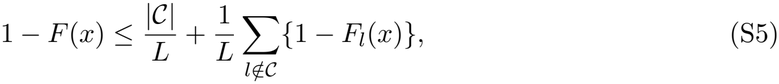

where the second term on the right-hand side of (S5) converges to 0 in probability. Since |*C*|*/L < α/*2, we have

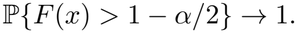

By the definition of *q*_1−*α/*2_, *F* (*x*) *>* 1 − *α/*2 implies *q*_1−*α/*2_ ≤ *x < µ*, i.e., *µ* ∉ *I*_eCI_(*α*). Therefore, we obtain

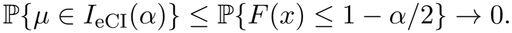

The case of 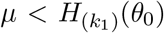 is proved in the same manner, where ℙ*{F* (*x*) *< α/*2*}* → 1 for 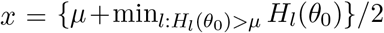, and by the definition of *q*_*α/*2_, *F* (*x*) *< α/*2 implies *q*_*α/*2_ ≥ *x > µ*. □

#### S3.3 Proof of Theorem 3.2

We prove Theorem 3.2 by using Lemmas S1 and S2.

*Proof of Theorem 3.2*. Since (i) follows from the argument for (ii) with *L* = 1, we first prove (ii). For *l* = 1, …, *L*, let *r*_*l*_ denote the rank of *H*_*l*_(*θ*_0_) among the *L* values *H*_1_(*θ*_0_), …, *H*_*L*_(*θ*_0_). Let

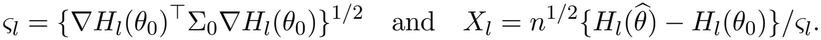

For *x* ∈ ℝ, let

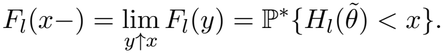

Since the convergence in Lemma S2(i) is uniform in *y* ∈ ℝ, from Lemma S2(i), we have

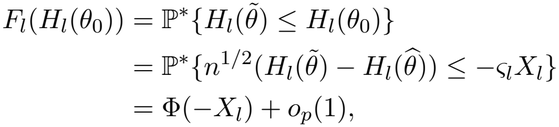

and similarly, from the counterpart with *<*, we have

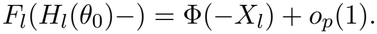

From Lemma S2(ii), −*X*_*l*_ converges in distribution to *N*(0, 1), and Φ(−*X*_*l*_) converges in distribution to the uniform distribution on (0, 1).

For *k ≠ l*, since *H*_1_(*θ*_0_), …, *H*_*L*_(*θ*_0_) are distinct, we have either *H*_*k*_(*θ*_0_) *< H*_*l*_(*θ*_0_), i.e., *r*_*k*_ *< r*_*l*_, or *H*_*k*_(*θ*_0_) *> H*_*l*_(*θ*_0_), i.e., *r*_*k*_ *> r*_*l*_. If *r*_*k*_ *< r*_*l*_, from Lemma S1 with *H* = *H*_*k*_ and *x* = *H*_*l*_(*θ*_0_), we have

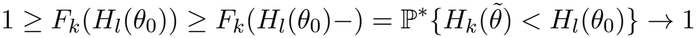

in probability. If *r*_*k*_ *> r*_*l*_, from Lemma S1 with *H* = *H*_*k*_ and *x* = *H*_*l*_(*θ*_0_), we have

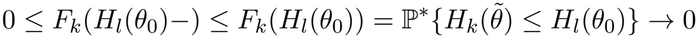

in probability. Since exactly *r*_*l*_ − 1 indices *k* satisfy *r*_*k*_ *< r*_*l*_, we obtain

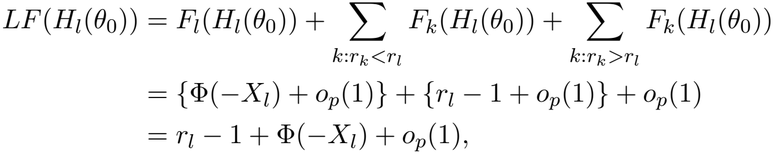

and in the same manner, we obtain

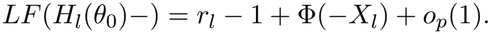

Let

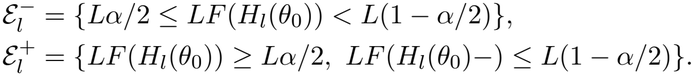

From (S2), we have

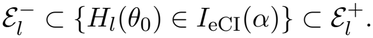

Let *ϑ* follow the uniform distribution on (0, 1), and let *λ*(*ℐ*) denote the length of an interval *ℐ* ⊂ ℝ. By Slutsky’s lemma and the portmanteau theorem (Lemmas 2.8 and 2.2 of van der Vaart, 1998), since ℙ*{r*_*l*_ − 1 + *ϑ* ∈ *{Lα/*2, *L*(1 − *α/*2)*}}* = 0, we have

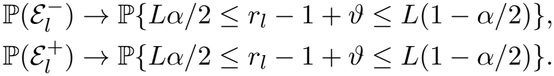

Since 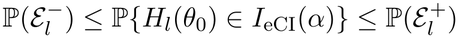, we obtain

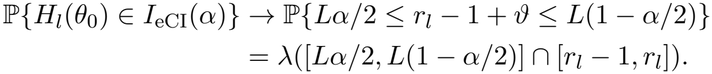

Since *s* is independent of the data and *r*_1_, …, *r*_*L*_ is a permutation of 1, …, *L*, we obtain

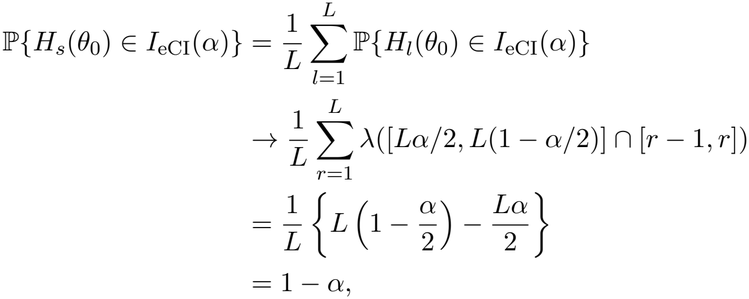

where we used 0 *< Lα/*2 *< L*(1 − *α/*2) *< L*.

We next prove (i). By the same argument as for (ii) with *L* = 1 and *H*_1_ replaced by *µ*_*L*_, which satisfies the conditions of Lemma S2 if Assumption 2 is satisfied and ∇*µ*_*L*_(*θ*_0_) *≠* 0, we obtain

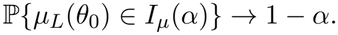

□

#### S3.4 Proof of Proposition 3.3

We prove Proposition 3.3 by using Lemma S1 and the argument in the proof of Theorem 3.1.

*Proof of Proposition 3.3*. We first prove (i). Suppose that *L <* 2*/α*. Given the GCMs, we consider the following three cases. If 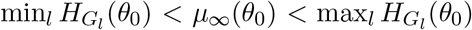, the argument in the proof of Theorem 3.1(i) with *µ* replaced by *µ*_∞_(*θ*_0_) and *k*_2_ = 1 shows that

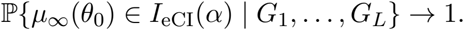

If 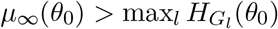, let 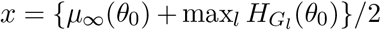. Then, from Lemma S1, we have *F* (*x*) → 1 in probability given the GCMs. By the definition of *q*_1−*α/*2_, *F* (*x*) *>* 1 − *α/*2 implies *q*_1−*α/*2_ ≤ *x < µ*_∞_(*θ*_0_), i.e., *µ*_∞_(*θ*_0_) ∉ *I*_eCI_(*α*). Therefore, we have

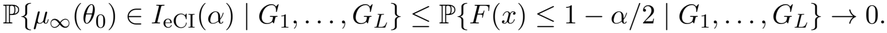

If 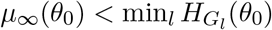, in the same manner, we have

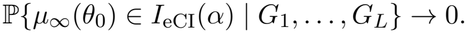

Since 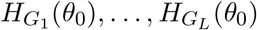 follow *N*(*µ*_∞_(*θ*_0_), *τ* ^2^), we have

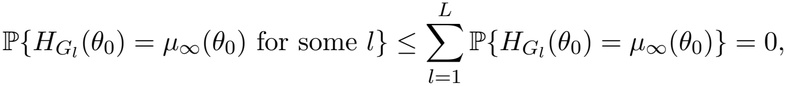

and hence, with probability one, 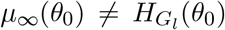 for *l* = 1, …, *L*, so that exactly one of 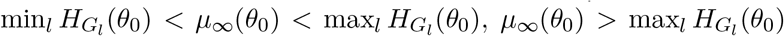, and 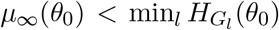 holds. Therefore, with probability one, we have

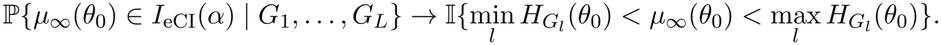

Since 0 ≤ ℙ*{µ*_∞_(*θ*_0_) ∈ *I*_eCI_(*α*) | *G*_1_, …, *G*_*L*_*}* ≤ 1, by the dominated convergence theorem, we obtain

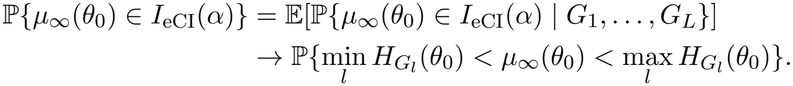

□

Since *H*_*G*_ (*θ*_0_), …, *H*_*G*_ (*θ*_0_) are independent and *µ*_∞_(*θ*_0_) is the median of *N*(*µ*_∞_(*θ*_0_), *τ* ^2^), we obtain

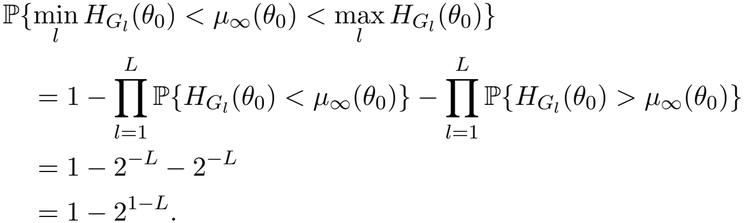

We next prove (ii). Let 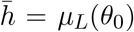. Given the GCMs, by Lemma S1 with *H* = *µ*_*L*_, both endpoints of *I*_*µ*_(*α*) converge to 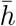 in probability, and hence, if 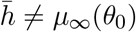, we have

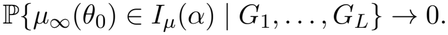

Since 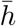 follows *N*(*µ*_∞_(*θ*_0_), *τ* ^2^*/L*), we have

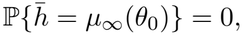

and hence, with probability one, we have

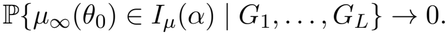

Since

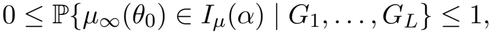

by the dominated convergence theorem, we obtain

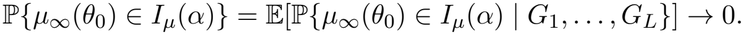

#### S3.5 Proof of Theorem 3.4

We prove Theorem 3.4 as follows.

*Proof of Theorem 3.4*. Let 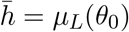 and

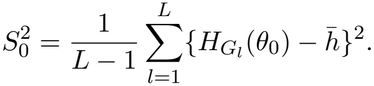

Since the data are independent of the GCMs, by Assumption 1 and the continuity of 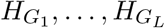 at *θ*_0_, given the GCMs, we have

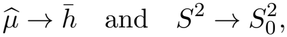

in probability. By the dominated convergence theorem, the same convergence holds under the joint distribution of the data and the GCMs. Let *U* follow the *t* distribution with *L* − 1 degrees of freedom.

We first prove (i). Since 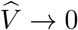 in probability and 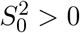 with probability one, we obtain

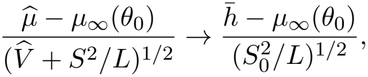

in probability. Since 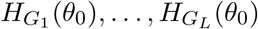 are independent and follow *N*(*µ*_∞_(*θ*_0_), *τ* ^2^),

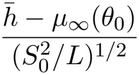

has the same distribution as *U*, which is continuous. From (2.3) of the main text, by Slutsky’s lemma and the portmanteau theorem, we obtain

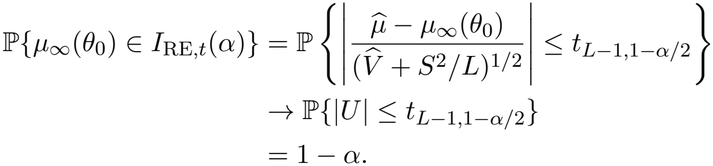

We next prove (ii). In the same manner as for (i), we have

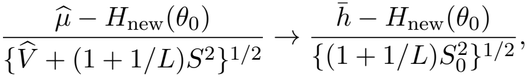

in probability. Since *H*_new_(*θ*_0_) follows *N*(*µ*_∞_(*θ*_0_), *τ* ^2^) independently of 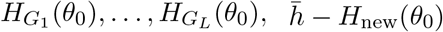 follows *N*(0, (1 + 1*/L*)*τ* ^2^) independently of 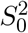, and hence

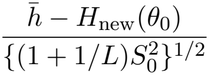

has the same distribution as *U* . From (2.4) of the main text, in the same manner as for (i), we obtain

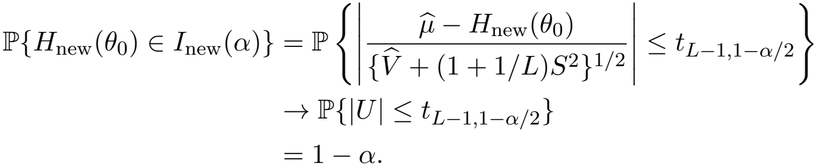

□

For *I*_RE,sim_ (*α*), given the data and the 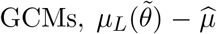 converges to 0 in probability by Lemma S1, and hence the endpoints of *I*_RE,sim_(*α*) satisfy the same convergence as those of *I*_RE,*t*_(*α*).

### S4 Location-shift model

#### S4.1 Exact coverage

For *e* = (*e*_1_, …, *e*_*L*_)^⊤^ ∈ ℝ^*L*^, define

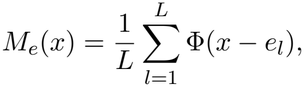

for *x* ∈ ℝ, and let 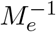 denote the inverse function of *M*_*e*_. Under model (3.1), let

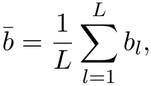

so that

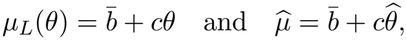

and let

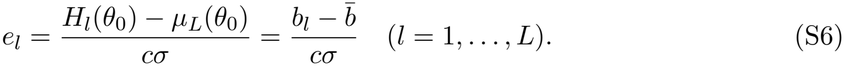

Let 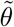 denote the coefficient sampled from *N*(*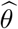, σ*^2^), and express it as 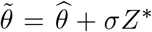, where *Z*^*^ follows *N*(0, 1) independently of the data. Since

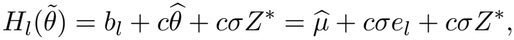

the cumulative distribution function of 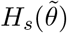 in Section S2 is given by, for *x* ∈ ℝ,

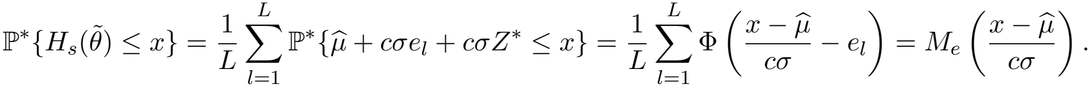

Since *M*_*e*_ is continuous and strictly increasing, condition (S1) holds, and the conditional *u*-quantile of 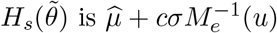. Thus, in the limit as *J* → ∞, we have

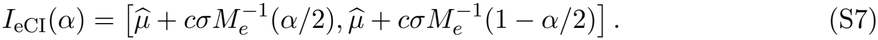

The interval *I*_eCI_(*α*) contains *µ*_*L*_(*θ*_0_) if and only if

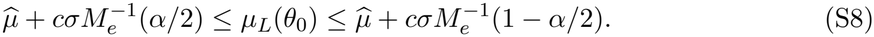

Since 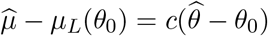, the inequalities in (S8) are equivalent to

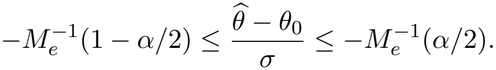

Because 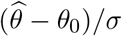 follows *N*(0, 1), the coverage of *I*_eCI_(*α*) is given by

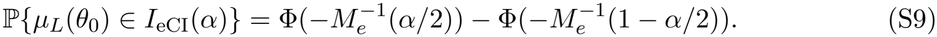

The coverage of *I*_eCI_(*α*) thus depends on the GCMs only through *e*.

Let *z*_1−*α/*2_ = Φ^−1^(1 − *α/*2). Regarding *I*_*µ*_(*α*), in the limit as *J* → ∞, we have

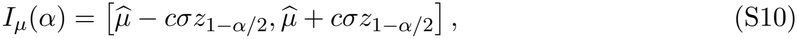

since

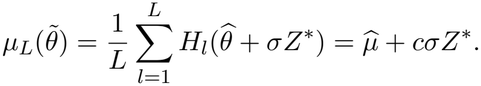

Thus, *I*_*µ*_(*α*) contains *µ*_*L*_(*θ*_0_) if and only if

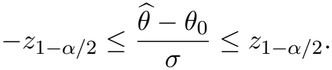

Therefore, the coverage of *I*_*µ*_(*α*) is given by

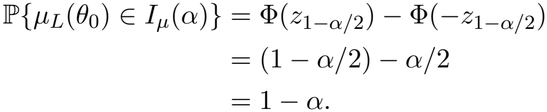

We next consider the predictand

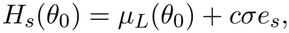

where *s* is uniformly distributed on *{*1, …, *L}* and independent of 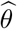. Since

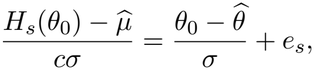

from (S7), *H*_*s*_(*θ*_0_) ∈ *I*_eCI_(*α*) holds if and only if

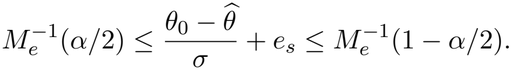

Since 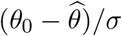 follows *N*(0, 1) and is independent of *s*, for *x* ∈ ℝ, we have

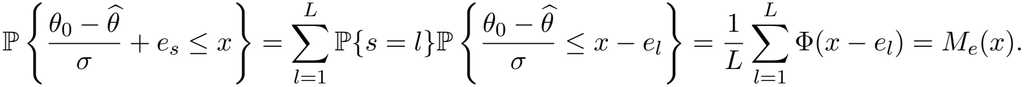

Thus, we obtain

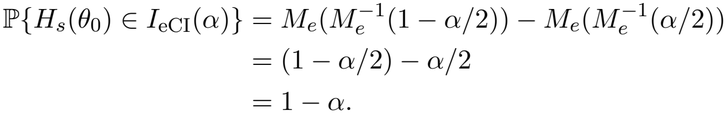

#### S4.2 Numerical examples

Table S2 shows the coverage (S9) at 1 − *α* = 0.95 for configurations, where we set *c* = *σ* = 1 and *θ*_0_ = 0. In the symmetric configurations with *L* = 3 and (*b*_1_, *b*_2_, *b*_3_) = (−*δ*, 0, *δ*), the coverage of *I*_eCI_(0.05) increased from 0.950 to 0.999 as *δ* increased from 0 to 2, and its width increased from 1.00 to 1.76 times that of *I*_*µ*_(0.05). In the configuration with *L* = 50, *b*_1_ = *· · ·* = *b*_49_ = 0, and *b*_50_ = 100, the coverage of *I*_eCI_(0.05) was 0.715, although its width was 1.15 times that of *I*_*µ*_(0.05). In this configuration, the distant GCM has the weight 1*/*50 = 0.02 in the mixture, which is smaller than the probability 0.025 excluded in the upper tail, and the upper endpoint of *I*_eCI_(0.05) is determined by the other 49 GCMs, whereas *µ*_*L*_(*θ*_0_) is shifted by 100*/*50 = 2 by the distant GCM.

**Table S2:** Exact coverage of *I*_*µ*_(0.05) and *I*_eCI_(0.05) for *µ*_*L*_(*θ*_0_) and the ratio of the width of *I*_eCI_(0.05) to that of *I*_*µ*_(0.05) under model (3.1) with *c* = *σ* = 1 and *θ*_0_ = 0. In the symmetric configurations, (*b*_1_, *b*_2_, *b*_3_) = (−*δ*, 0, *δ*). In the configuration with one distant GCM, *b*_1_ = *· · ·* = *b*_49_ = 0 and *b*_50_ = 100.

| Configuration | *L* | *μ*<sub>*L*</sub>(*θ*<sub>0</sub>) | Coverage of *I*<sub>*μ*</sub> | Coverage of *I*<sub>eCI</sub> | Width ratio |
|---|---|---|---|---|---|
| Symmetric, *δ* = 0 | 3 | 0 | 0.950 | 0.950 | 1.000 |
| Symmetric, *δ* = 0.5 | 3 | 0 | 0.950 | 0.965 | 1.079 |
| Symmetric, *δ* = 1 | 3 | 0 | 0.950 | 0.987 | 1.270 |
| Symmetric, *δ* = 2 | 3 | 0 | 0.950 | 0.999 | 1.756 |
| One distant GCM | 50 | 2 | 0.950 | 0.715 | 1.153 |

#### S4.3 Normal random-effects model

In the normal random-effects model, *b*_1_, …, *b*_*L*_ in model (3.1) are independently distributed as *N*(*b*_0_, *τ* ^2^) and independent of *θ*. Then, we have *µ*_∞_(*θ*_0_) = *b*_0_ + *cθ*_0_, *V* = *c*^2^*σ*^2^, and 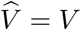 for *J* → ∞. Since

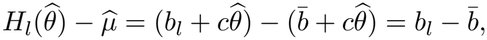

we have

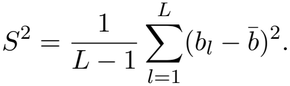

We also have

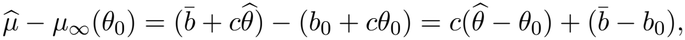

which follows *N*(0, *V* + *τ* ^2^*/L*) and is independent of *S*^2^, since 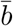 and *S*^2^ are independent for the normal sample *b*_1_, …, *b*_*L*_ and 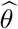 is independent of *b*_1_, …, *b*_*L*_. Let

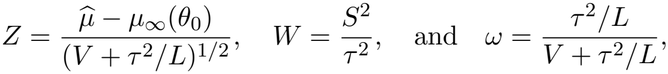

where *Z* follows *N*(0, 1), (*L* − 1)*W* follows the chi-squared distribution with *L* − 1 degrees of freedom, and *Z* and *W* are independent. Since

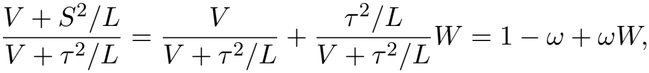

from (2.3) of the main text, we have

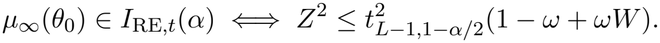

Therefore, by the independence of *Z* and *W*, we obtain

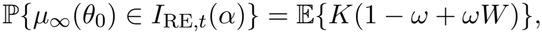

where 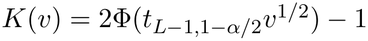 for *v* ≥ 0.

Similarly, let *b*_*L*+1_ follow *N*(*b*_0_, *τ* ^2^) independently of *b*_1_, …, *b*_*L*_ and 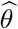. Then, 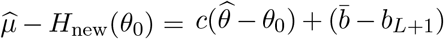 follows *N*(0, *V* + (1 + 1*/L*)*τ* ^2^) and is independent of *S*^2^. From (2.4) of the main text, in the same manner, we obtain

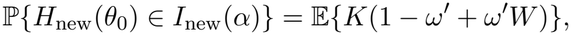

where *ω*^*′*^ = (1 + 1*/L*)*τ* ^2^*/{V* + (1 + 1*/L*)*τ* ^2^*}*.

Since 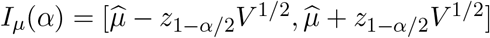 from (S10), we have

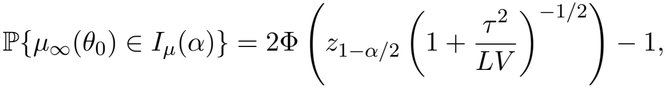

which is smaller than 1 − *α* for *τ* ^2^ *>* 0.

For *I*_RE,sim_(*α*), in the limit as *J* → ∞, the endpoints are the *α/*2- and (1 − *α/*2)-quantiles of 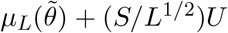 given the data, where *U* follows the *t* distribution with *L* − 1 degrees of freedom independently of *Z*^*^. Since 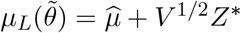 from Section S4.1, we have

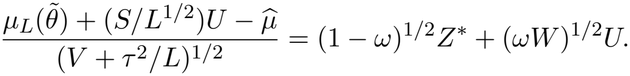

Let *q*_RE_(*W*) denote the (1 − *α/*2)-quantile of (1 − *ω*)^1*/*2^*Z*^*^ + (*ωW*)^1*/*2^*U* given *W* . Since the conditional distribution of (1 − *ω*)^1*/*2^*Z*^*^ + (*ωW*)^1*/*2^*U* given *W* is symmetric about 0, we have

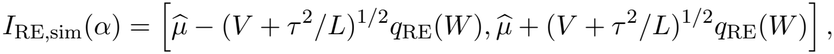

and hence *µ*_∞_(*θ*_0_) ∈ *I*_RE,sim_(*α*) if and only if |*Z*| ≤ *q*_RE_(*W*). Therefore, by the independence of *Z* and *W*, we obtain

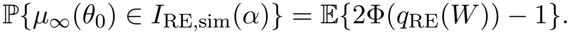

For *I*_eCI_(*α*), by the same derivation as for (S9), we have

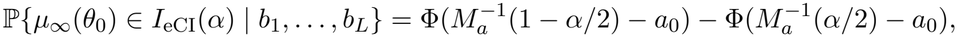

where *a* = (*a*_1_, …, *a*_*L*_)^⊤^ with *a*_*l*_ = *b*_*l*_*/*(*cσ*) and *a*_0_ = *b*_0_*/*(*cσ*).

To illustrate these expressions, we evaluated the coverage at 1−*α* = 0.95 for *L* ∈ *{*3, 5, 10, 20*}* and 0.01 ≤ *τ* ^2^*/V* ≤ 100. We computed E*{K*(1 − *ω* + *ωW*)*}* and E*{K*(1 − *ω*^*′*^ + *ω*^*′*^*W*)*}* by the midpoint rule on 20,000 quantiles of *W*, E*{*2Φ(*q*_RE_(*W*)) − 1*}* by the midpoint rule on 400 quantiles of *W* and 2,000 quantiles of *U*, and the coverage of *I*_eCI_(*α*) by averaging ℙ*{µ*_∞_(*θ*_0_) ∈ *I*_eCI_(*α*) | *b*_1_, …, *b*_*L*_*}* over 20,000 draws of *b*_1_, …, *b*_*L*_.

Figure S1 shows the results. For *L* ∈ *{*3, 5, 10, 20*}* and 0.01 ≤ *τ* ^2^*/V* ≤ 100, the coverage of *I*_RE,*t*_(0.05) for *µ*_∞_(*θ*_0_) was at least 0.954, and that of *I*_new_(0.05) for *H*_new_(*θ*_0_) was at least 0.950.

**Figure S1:**
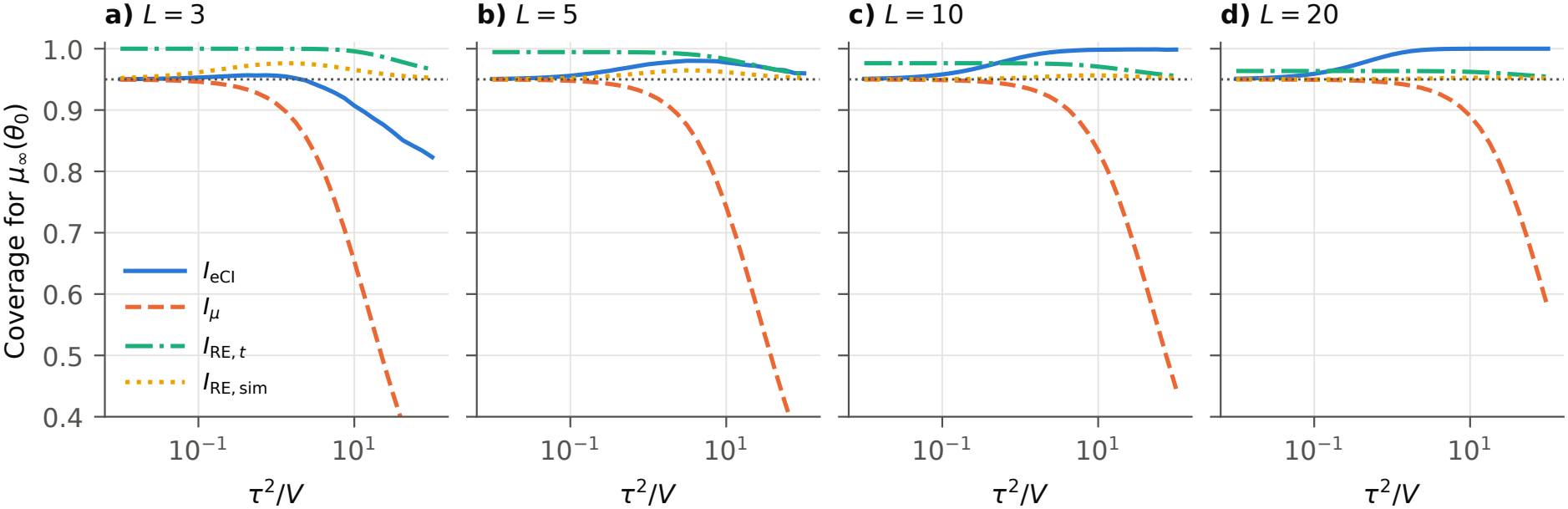
Coverage of *I*_eCI_(0.05), *I*_*µ*_(0.05), *I*_RE,*t*_(0.05), and *I*_RE,sim_(0.05) for *µ*_∞_(*θ*_0_) under the normal random-effects model as functions of *τ* ^2^*/V* on a logarithmic scale. The dotted lines are at 0.95.

### S5 Details of the simulation study

#### S5.1 Data generation and computation

The coefficient vector *θ*_0_ was computed by least squares on 1,000 equally spaced temperatures between the boundary knots. The knots of the natural cubic spline basis were the percentiles of a temperature series of 500 years generated in the same manner as the observed temperature, and they were −1.38, 4.55, 15.75, 18.41, and 24.34. The basis was computed for the scaled temperature (*T* − 15)*/*10, and it is linear below the lower boundary knot and above the upper boundary knot. In the computation of *H*_*l*_(*θ*), we rounded the temperature down to bins of width 0.1 ^°^C and evaluated *f*_*θ*_ at the midpoints of the bins, and *H*_*l*_(*θ*_0_) was computed in the same manner. In the large-ensemble design, the *L* − 1 members with Δ_*l*_ = 2.5 had temperature series generated independently, so that the members differ only in the internal variability.

#### S5.2 Additional results

Tables S3–S5 show the coverage for the fixed ensembles with *D* = 20, the coverage of *I*_eCI_(*α*) for *H*_*s*_(*θ*_0_), and the results for the random ensembles at 1 − *α* = 0.66. With *D* = 20, the coverage of *I*_*µ*_(*α*) was between 0.941 and 0.944 at 1 − *α* = 0.95 and between 0.660 and 0.664 at 1 − *α* = 0.66. For the random ensembles at 1 − *α* = 0.66, the coverage for *µ*_∞_(*θ*_0_) was at least 0.676 for *I*_RE,*t*_(0.34), between 0.659 and 0.699 for *I*_RE,sim_(0.34), between 0.704 and 0.922 for *I*_eCI_(0.34), and between 0.453 and 0.644 for *I*_*µ*_(0.34), and the coverage for *H*_new_(*θ*_0_) was between 0.539 and 0.661 for *I*_eCI_(0.34) and at least 0.680 for *I*_new_(0.34).

**Table S3:**
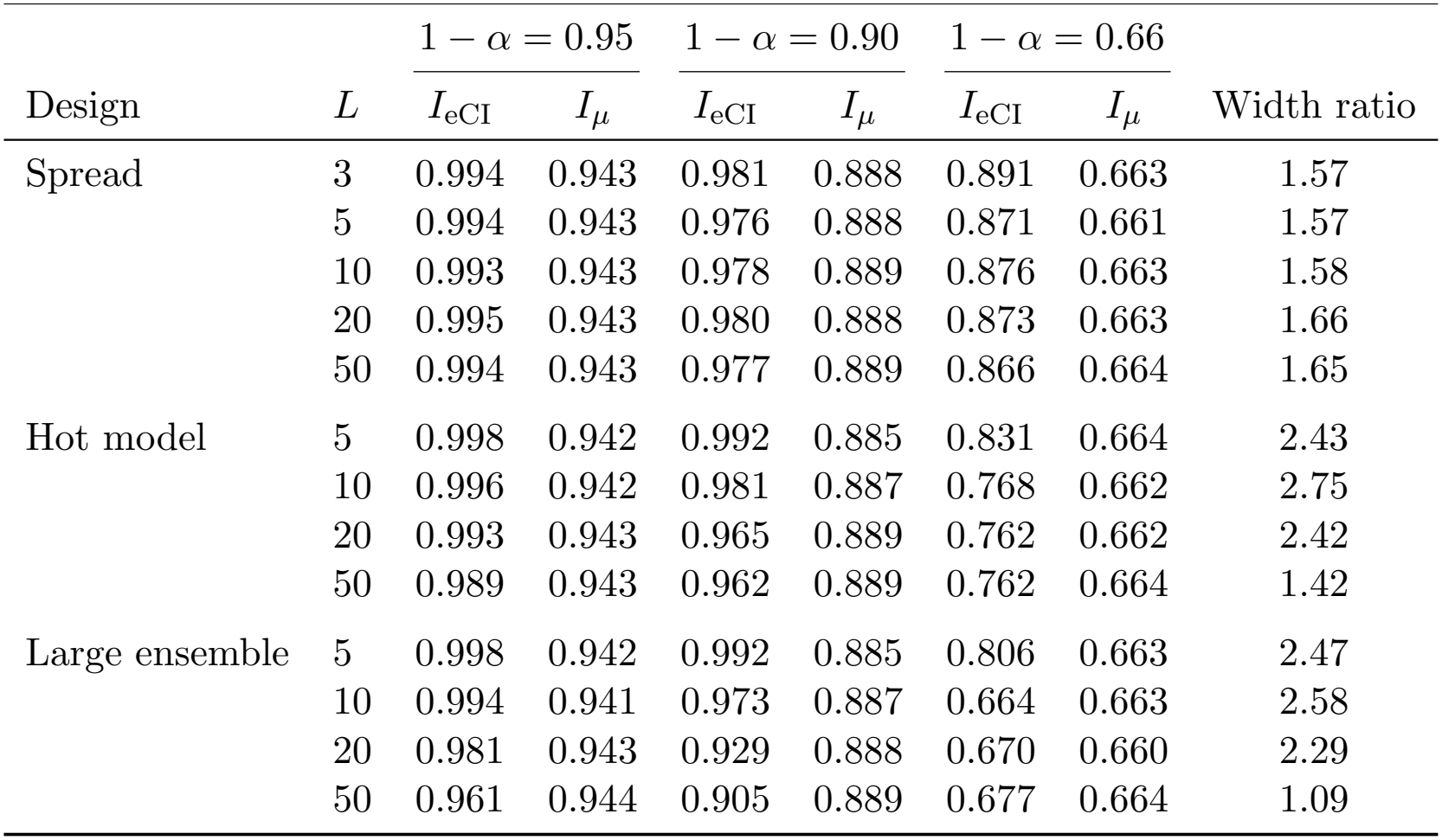
Coverage of *I*_eCI_(*α*) and *I*_*µ*_(*α*) for *µ*_*L*_(*θ*_0_) in the fixed ensembles with *D* = 20, and the ratio of the median width of *I*_eCI_(0.05) to that of *I*_*µ*_(0.05).

| Design | *L* | 1 − α = 0.95 |  | 1 − α = 0.90 |  | 1 − α = 0.66 |  | Width ratio |
|---|---|---|---|---|---|---|---|---|
|  |  | *I*<sub>eCI</sub> | *I*<sub>μ</sub> | *I*<sub>eCI</sub> | *I*<sub>μ</sub> | *I*<sub>eCI</sub> | *I*<sub>μ</sub> |  |
| Spread | 3 | 0.994 | 0.943 | 0.981 | 0.888 | 0.891 | 0.663 | 1.57 |
|  | 5 | 0.994 | 0.943 | 0.976 | 0.888 | 0.871 | 0.661 | 1.57 |
|  | 10 | 0.993 | 0.943 | 0.978 | 0.889 | 0.876 | 0.663 | 1.58 |
|  | 20 | 0.995 | 0.943 | 0.980 | 0.888 | 0.873 | 0.663 | 1.66 |
|  | 50 | 0.994 | 0.943 | 0.977 | 0.889 | 0.866 | 0.664 | 1.65 |
| Hot model | 5 | 0.998 | 0.942 | 0.992 | 0.885 | 0.831 | 0.664 | 2.43 |
|  | 10 | 0.996 | 0.942 | 0.981 | 0.887 | 0.768 | 0.662 | 2.75 |
|  | 20 | 0.993 | 0.943 | 0.965 | 0.889 | 0.762 | 0.662 | 2.42 |
|  | 50 | 0.989 | 0.943 | 0.962 | 0.889 | 0.762 | 0.664 | 1.42 |
| Large ensemble | 5 | 0.998 | 0.942 | 0.992 | 0.885 | 0.806 | 0.663 | 2.47 |
|  | 10 | 0.994 | 0.941 | 0.973 | 0.887 | 0.664 | 0.663 | 2.58 |
|  | 20 | 0.981 | 0.943 | 0.929 | 0.888 | 0.670 | 0.660 | 2.29 |
|  | 50 | 0.961 | 0.944 | 0.905 | 0.889 | 0.677 | 0.664 | 1.09 |

### S6 Details of the reanalysis

Table S6 shows 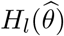, the percentile interval, and the standard deviation of *H*_*l*_(*θ*^(*j*)^) over the *J* = 100 draws for each GCM, and the share of the variance among the GCMs in the variance of the pooled impacts.

**Table S4:**
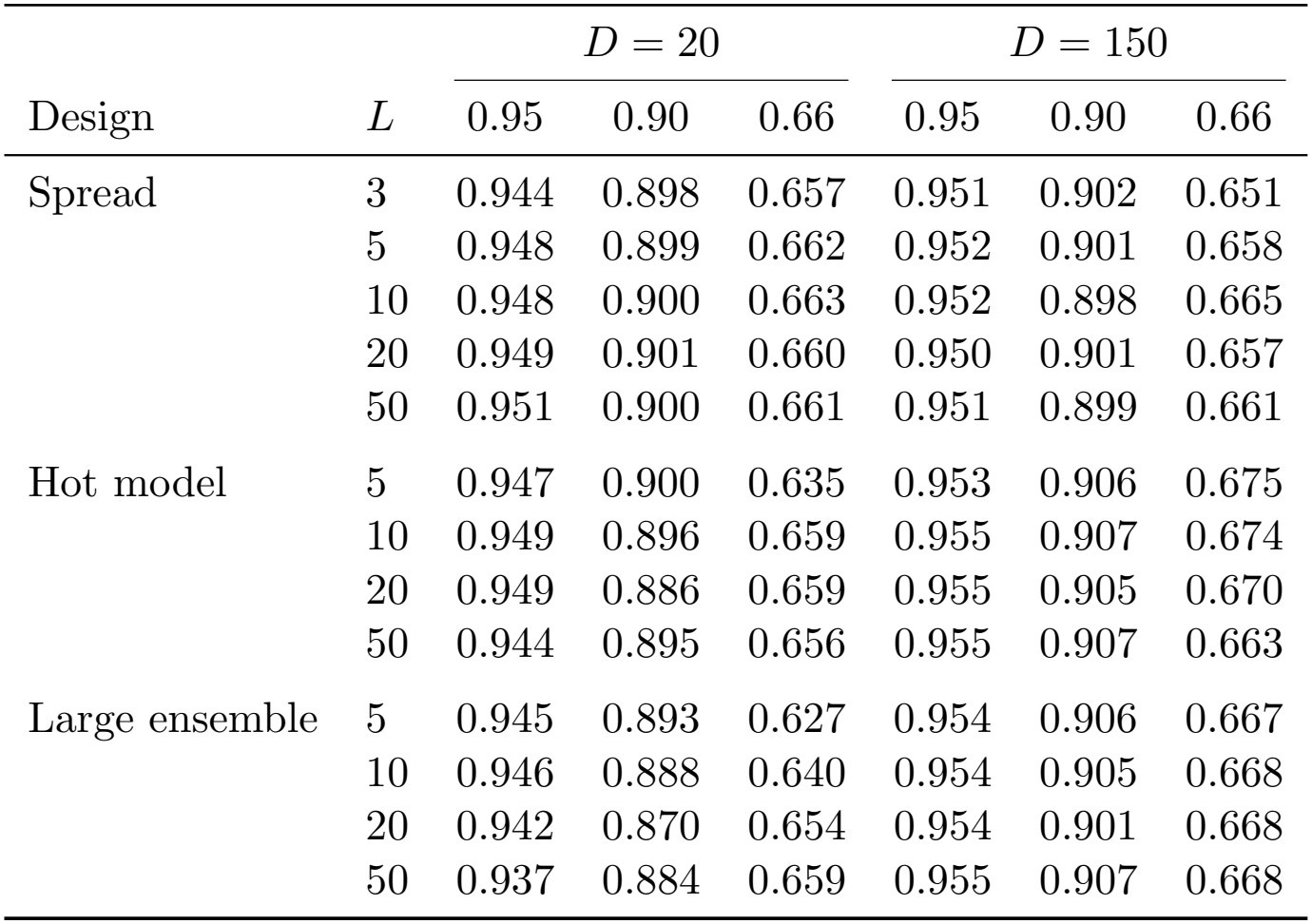
Coverage of *I*_eCI_(*α*) for *H*_*s*_(*θ*_0_) in the fixed ensembles at 1 − *α* ∈ *{*0.95, 0.90, 0.66*}*.

| Design | *L* | *D* = 20 |  |  | *D* = 150 |  |  |
|---|---|---|---|---|---|---|---|
|  |  | 0.95 | 0.90 | 0.66 | 0.95 | 0.90 | 0.66 |
| Spread | 3 | 0.944 | 0.898 | 0.657 | 0.951 | 0.902 | 0.651 |
|  | 5 | 0.948 | 0.899 | 0.662 | 0.952 | 0.901 | 0.658 |
|  | 10 | 0.948 | 0.900 | 0.663 | 0.952 | 0.898 | 0.665 |
|  | 20 | 0.949 | 0.901 | 0.660 | 0.950 | 0.901 | 0.657 |
|  | 50 | 0.951 | 0.900 | 0.661 | 0.951 | 0.899 | 0.661 |
| Hot model | 5 | 0.947 | 0.900 | 0.635 | 0.953 | 0.906 | 0.675 |
|  | 10 | 0.949 | 0.896 | 0.659 | 0.955 | 0.907 | 0.674 |
|  | 20 | 0.949 | 0.886 | 0.659 | 0.955 | 0.905 | 0.670 |
|  | 50 | 0.944 | 0.895 | 0.656 | 0.955 | 0.907 | 0.663 |
| Large ensemble | 5 | 0.945 | 0.893 | 0.627 | 0.954 | 0.906 | 0.667 |
|  | 10 | 0.946 | 0.888 | 0.640 | 0.954 | 0.905 | 0.668 |
|  | 20 | 0.942 | 0.870 | 0.654 | 0.954 | 0.901 | 0.668 |
|  | 50 | 0.937 | 0.884 | 0.659 | 0.955 | 0.907 | 0.668 |

**Table S5:** Coverage of the intervals at 1 − *α* = 0.66 for *µ*_∞_(*θ*_0_) and *H*_new_(*θ*_0_) in the random ensembles, and the ratio of the median width of *I*_RE,*t*_(0.34) to that of *I*_eCI_(0.34).

|  |  | Coverage for *μ*<sub>∞</sub>(*θ*<sub>0</sub>) |  |  |  | Coverage for *H*<sub>new</sub>(*θ*<sub>0</sub>) |  |  |
|---|---|---|---|---|---|---|---|---|
| *D* | *L* | *I*<sub>eCI</sub> | *I*<sub>*μ*</sub> | *I*<sub>RE,*t*</sub> | *I*<sub>RE,sim</sub> | *I*<sub>eCI</sub> | *I*<sub>new</sub> | Width ratio |
| 20 | 3 | 0.704 | 0.524 | 0.748 | 0.693 | 0.581 | 0.741 | 1.13 |
|  | 5 | 0.777 | 0.575 | 0.708 | 0.675 | 0.609 | 0.707 | 0.87 |
|  | 10 | 0.812 | 0.611 | 0.680 | 0.659 | 0.625 | 0.681 | 0.73 |
|  | 20 | 0.858 | 0.644 | 0.681 | 0.667 | 0.647 | 0.680 | 0.68 |
| 150 | 3 | 0.728 | 0.453 | 0.728 | 0.699 | 0.539 | 0.719 | 0.99 |
|  | 5 | 0.833 | 0.504 | 0.699 | 0.680 | 0.598 | 0.686 | 0.72 |
|  | 10 | 0.879 | 0.545 | 0.676 | 0.662 | 0.630 | 0.685 | 0.58 |
|  | 20 | 0.922 | 0.614 | 0.677 | 0.665 | 0.661 | 0.694 | 0.51 |

**Table S6:**
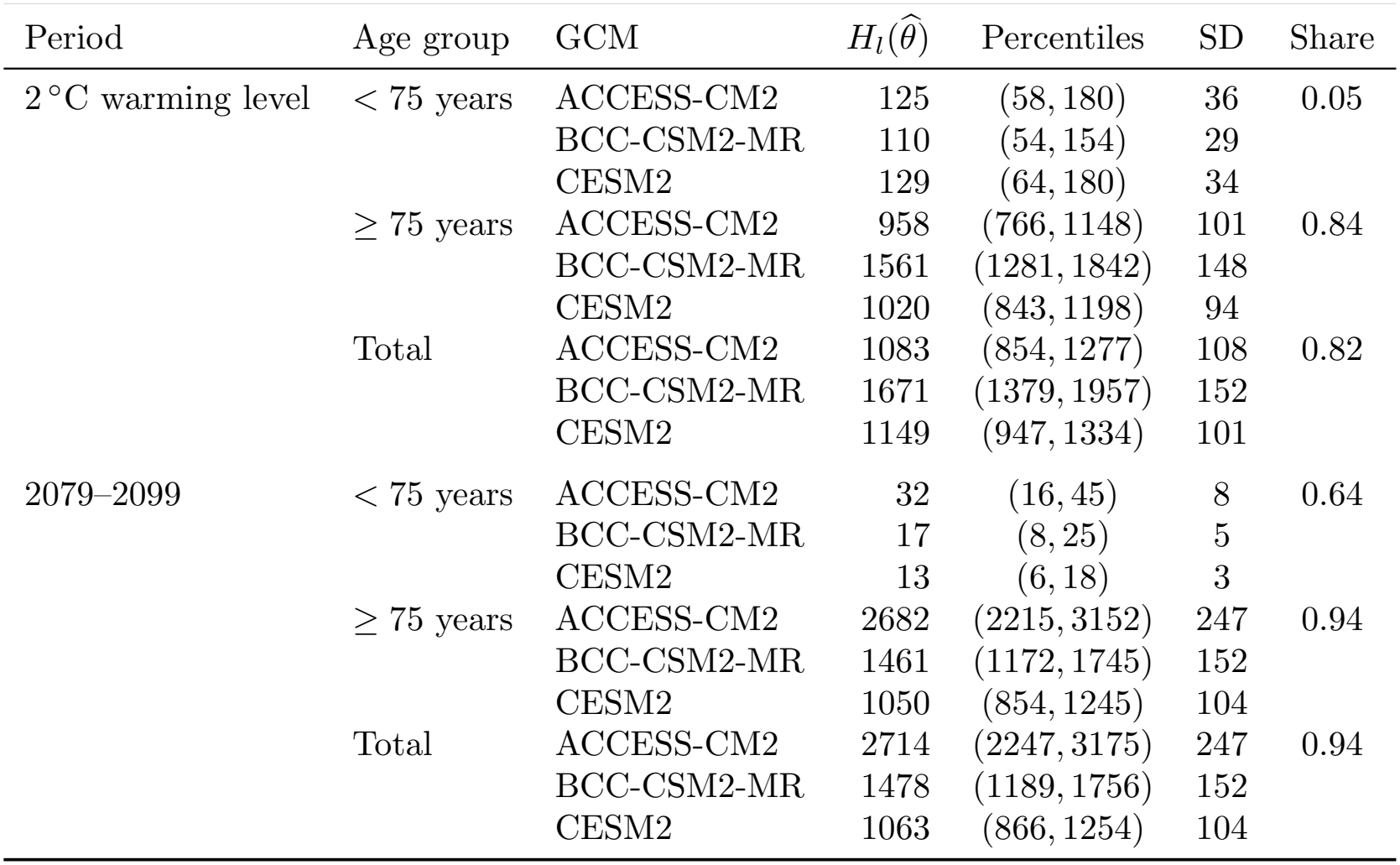
Annual heat-related deaths in London under each GCM in the two 21-year periods: 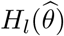, the 2.5th and 97.5th percentiles and the standard deviation (SD) of *H*_*l*_(*θ*^(*j*)^) over the draws, and the share of the variance among the GCMs in the variance of the pooled impacts.

| Period | Age group | GCM | H<sub>l</sub>(θ̂) | Percentiles | SD | Share |
|---|---|---|---|---|---|---|
| 2 °C warming level | < 75 years | ACCESS-CM2 | 125 | (58, 180) | 36 | 0.05 |
|  |  | BCC-CSM2-MR | 110 | (54, 154) | 29 |  |
|  |  | CESM2 | 129 | (64, 180) | 34 |  |
|  | ≥ 75 years | ACCESS-CM2 | 958 | (766, 1148) | 101 | 0.84 |
|  |  | BCC-CSM2-MR | 1561 | (1281, 1842) | 148 |  |
|  |  | CESM2 | 1020 | (843, 1198) | 94 |  |
|  | Total | ACCESS-CM2 | 1083 | (854, 1277) | 108 | 0.82 |
|  |  | BCC-CSM2-MR | 1671 | (1379, 1957) | 152 |  |
|  |  | CESM2 | 1149 | (947, 1334) | 101 |  |
| 2079–2099 | < 75 years | ACCESS-CM2 | 32 | (16, 45) | 8 | 0.64 |
|  |  | BCC-CSM2-MR | 17 | (8, 25) | 5 |  |
|  |  | CESM2 | 13 | (6, 18) | 3 |  |
|  | ≥ 75 years | ACCESS-CM2 | 2682 | (2215, 3152) | 247 | 0.94 |
|  |  | BCC-CSM2-MR | 1461 | (1172, 1745) | 152 |  |
|  |  | CESM2 | 1050 | (854, 1245) | 104 |  |
|  | Total | ACCESS-CM2 | 2714 | (2247, 3175) | 247 | 0.94 |
|  |  | BCC-CSM2-MR | 1478 | (1189, 1756) | 152 |  |
|  |  | CESM2 | 1063 | (866, 1254) | 104 |  |

